# Salience Network Connectivity Relates to Sleep and Sensory Over-Responsivity in Infants at High and Low Likelihood for Autism

**DOI:** 10.64898/2026.01.13.26344039

**Authors:** Emily Chiem, Lauren Wagner, Leanna M. Hernandez, Shulamite Green, Mirella Dapretto

## Abstract

Sleep problems and sensory over-responsivity (SOR) are common, co-occurring, and early-emerging features of Autism Spectrum Disorder (ASD). Yet, the early neural mechanisms underlying this relationship remain unclear. Here, we used resting-state fMRI data from the Infant Brain Imaging Study (IBIS) to examine how brain connectivity at 6 months may relate to parent-reported measures of sleep-onset problems and SOR in infants at varying familial likelihood for ASD. The right anterior insula was used in seed-based analyses to investigate Salience Network (SN) connectivity to cortical and cerebellar regions of interest previously implicated in sleep disruption, sensory processing challenges, and ASD. Infants at high (HL) and low (LL) likelihood for ASD displayed divergent patterns of SN connectivity with sensorimotor cortex, as well as cerebellar regions involved in sensorimotor processing and higher-order functions. Furthermore, stronger SN connectivity with sensorimotor cortices and cerebellar regions was associated with worse sleep-onset problems and SOR in HL infants. In contrast, stronger SN-cerebellar connectivity was related to fewer sleep-onset problems and SOR in LL infants. Our findings indicate that altered SN connectivity may result in over-attribution of attention to sensory stimuli and highlight aberrant sensory prediction learning, which may underlie worse sleep problems and higher SOR in HL infants.

## Introduction

Sleep problems are pervasive in Autism Spectrum Disorder (ASD), occurring in up to 80% of affected children [1] and are some of the first concerns raised by parents prior to a formal diagnosis [2]. While the core diagnostic criteria for ASD include social-communication difficulties and restrictive, repetitive behaviors [3], sleep disruption is another common feature of ASD that persists throughout the lifespan with profound consequences on cognition and overall daily functioning [4], [5]. Prior work has shown that sleep disruption in infancy has negative effects on developmental outcomes, such as cognition [6], [7], [8], language [9], [10], motor skills [11], and even later ASD symptomatology [12], [13]. Yet, our understanding of the impact of sleep problems on neurodevelopment, particularly in the first few years of life, remains limited. To our knowledge, only one study has investigated how infant sleep disturbances relate to brain development in ASD [14]. This study showed that difficulties with sleep initiation are associated with reduced subcortical brain volume in infants who later received an ASD diagnosis. Despite the high prevalence of sleep problems in autism and their potential to impact later developmental outcomes, the relationship between sleep problems in infancy and functional brain connectivity in ASD remains wholly uncharted.

Sensory processing challenges were added as a core symptom of ASD in the DSM-V [3] and can emerge early amongst autistic individuals. These sensory atypicalities often manifest as sensory over-responsivity (SOR) – characterized by extreme responses to or avoidance of sensory stimuli [15], [16]. This pattern is thought to emerge as early as infancy, prior to clinical diagnosis of ASD [17]. Past work has also shown that autistic toddlers display increased SOR compared to toddlers without a diagnosis [18], [19], [20] and that these sensory atypicalities are negatively associated with later adaptive behavior [21]. Notably, several studies have demonstrated a significant co-occurring, positive association between sleep problems and sensory processing differences in children and adolescents with ASD [22], [23], [24], [25], [26], [27], such that worse sleep problems are related to increased sensory responsivity. Furthermore, both sleep problems and sensory challenges are related to worse ASD symptomatology, such as social functioning [28], [29] and repetitive behaviors [30], [31]. The correlation between autism-associated sleep problems and sensory atypicalities consistently reported from toddlerhood to adolescence suggests that these symptoms may interact throughout development.

Although the co-occurrence of sleep and sensory processing challenges in ASD has been well-documented, the neural mechanisms underlying these associations have yet to be elucidated. It also remains unclear how early autism-related differences in brain connectivity manifest, or how they may influence the relationship between sleep problems and sensory processing atypicalities in ASD. Toddlers are typically among the youngest age groups investigated in sleep and sensory studies of ASD [15], [24], despite the fact that the biological bases for sleep and sensory problems likely goes awry during the fetal and/or neonatal period. While several theories have been proposed to explain the relationship between sleep disturbances and sensory responsivity in ASD, the underlying mechanisms are likely complex and multifactorial. Some have suggested that poor sleep may exacerbate core ASD symptoms, such as difficulty regulating sensory inputs [32], [33]. On the other hand, because the sensory gating process largely modulates sleep [34], the inability to properly attenuate sensory inputs may disrupt sleep onset and sleep maintenance [35]. Given the intertwined and reciprocal nature of sleep and sensory processes, early perturbations in their potentially shared neural mechanisms may exacerbate these symptoms and contribute to altered developmental trajectories.

The brain’s functional connectome emerges *in utero* [36]. Large-scale functional connectivity networks undergo dramatic development during infancy, with the salience network (SN) – a functional network involved in detecting and prioritizing relevant stimuli [37] – amongst the first to mature [38]. SN connectivity is consistently altered in individuals with ASD. For instance, hyperconnectivity between core SN nodes has been observed in autistic children and was related to more severe restricted and repetitive behaviors [39]. Other work in autistic adults has implicated both hypo- and hyperconnectivity of the SN, including decreased connectivity between the prefrontal cortex and cerebellum, and increased connectivity between temporoparietal regions [40]. Even earlier in development, recent work has shown that stronger connectivity between the salience and visual networks was associated with higher levels of social impairment in school-aged children at varying genetic liability for ASD [41]. Importantly, stronger SN connectivity with the sensorimotor cortices is related to worse sensory over-responsivity in autistic youth [42], [43]. Interestingly, this association between stronger SN connectivity with sensory processing regions and sensory over-responsivity, previously reported in older individuals [42], [43] is present as early as 6 weeks in infants at high likelihood for ASD [44], suggesting that an over-attribution of attention to extraneous sensory stimuli is present quite early in development.

While three studies to date have investigated SN connectivity in infants at increased familial likelihood for ASD [44], [45], [46], the relationships between SN connectivity, sleep problems, and sensory challenges in this infant population have yet to be investigated. Since 20% of infants who have an older sibling with ASD ultimately receive a diagnosis [47], prospective studies in this population provide a unique opportunity to examine how early functional brain atypicalities may predict later symptomatology and diagnostic outcomes. Indeed, several studies have shown early structural [48], [49] and functional [50], [51], [52], [53] brain differences in these infant siblings, which predicted later ASD symptomatology [54], [55], [56], [57], [58], [59]. Furthermore, parents of infants at high familial likelihood for ASD report greater sleep problems, such as longer sleep latency and decreased nighttime sleep duration [60]. Recent work has shown that 6-week-old infants at high familial likelihood for ASD exhibit increased SN connectivity to sensorimotor regions – which is predictive of later sensory sensitivities – as well as decreased connectivity to prefrontal regions in comparison to infants with no familial predisposition for ASD [44]. This altered SN connectivity, which suggests impairments in detecting and orienting towards salient stimuli, supports the hypothesis that autistic individuals may perceive low-level sensory inputs as more salient than socially relevant information [61]. Together, these findings suggest that differences in salience detection develops early in life.

In addition to these changes in SN connectivity, ASD has also been associated with altered cerebellar functional connectivity [62], [63], [64], [65], [66], [67]. Previous work has shown that children and adolescents with ASD show greater cerebellar functional connectivity with cortical sensorimotor networks [62] and that connectivity between these regions is associated with SOR symptoms [67]. In the context of a sparse, but growing, literature on the role of the cerebellum in infant behavioral development [68], very few studies (e.g., [69], [70], [71]) have focused on functional cerebellar connectivity, particularly in ASD. Functional cerebellar networks are present soon after birth [72] and cerebro-cerebellar networks have been defined in neonates [71] and 6-month-old infants [73]. Although studies of cerebellar functional connectivity in infants at familial likelihood for ASD are sparse, one prior study in 9-month-olds found that weaker connectivity between the right cerebellar crus I—implicated in language processing and working memory [74], [75] — and the frontal cortex was associated with delayed language trajectories [69] in infants at heightened and low likelihood for developing ASD.

While SN and cerebellar functional connectivity have independently been associated with ASD and related symptomatology, as well as with sleep disturbances [76], [77], [78], [79], [80], no study to our knowledge has leveraged these converging neural mechanisms to investigate the relationship between sleep and sensory processing problems in ASD. Here, we used functional magnetic resonance imaging (fMRI) to examine the early neural correlates of sleep problems and SOR in infants at high (HL; older sibling with a diagnosis) and low (LL; no family history) likelihood for ASD. The right anterior insula (rAI) – a main hub of the salience network – was used in seed-based analyses to examine how SN connectivity with cortical and cerebellar regions of interest at 6 months of age may relate to emerging sleep-onset problems and SOR symptoms. Based on prior literature showing atypical brain connectivity in HL infants and ASD youth, our *a priori* regions of interest included sensorimotor cortices [42], [44] and regions of the cerebellum involved in sensorimotor processing (lobules I-IV, lobules V-VI, lobules VIIIA/B; [67]) and higher-order functions (crus I; [69]). In addition, we examined how sleep-onset problems and SOR symptoms may be associated with later ASD-related symptomatology.

## Methods

### Participants

Participants were enrolled as part of the Infant Brain Imaging Study (IBIS). Publicly available data was downloaded from the National Institute of Mental Health Data Archive (Collection #19 and #2027). The IBIS a longitudinal study aimed at investigating early brain and behavioral development in infants at varying familial likelihood for ASD. High likelihood (HL) infants had at least one older sibling with a confirmed ASD diagnosis. Low likelihood (LL) infants had at least one older typically developing sibling and no first- or second-degree relatives with ASD or an intellectual disability. Parents provided written informed consent, and infants were screened and assessed at each data collection site: University of North Carolina at Chapel Hill, University of Washington, Children’s Hospital of Philadelphia, or Washington University in St Louis. Research protocols were approved by the institutional review boards at each site. Participants were excluded based on the following criteria: premature birth (gestational age < 36 weeks) or low birth weight (< 2,000g); significant medical or neurological conditions affecting growth, development, or cognition; maternal substance use during pregnancy; a first degree relative with psychosis, schizophrenia, or bipolar disorder; contraindication for MRI; non-English-speaking family; adoption; and twin birth [57].

A total of 101 infants (62 HL, 39 LL) with fMRI scans and Infant Behavior Questionnaire (IBQ) data, both collected at 6 months of age, were included in the present study. Demographics are summarized in Table 1. Groups were matched on sex, age at scan, race, ethnicity, and maternal education. Seventy-nine infants also had IBQ data collected at 12 months. Data from the Sensory Experiences Questionnaire (SEQ) was available at 12 months (N=76) and at 24 months (N=63). Eighty-eight infants received a clinical best-estimate diagnosis of ASD at 24 or 36 months based on the Diagnostic and Statistical Manual of Mental Disorders, fourth edition, text revision (DSM-IV-TR) criteria using all available testing and interview data including the Autism Diagnostic Observation Schedule (ADOS; [81]), the Autism Diagnostic Interview-Revised (ADI-R; [82]), and the Mullen Scales of Early Learning [83]. In the present HL sample, only 7 infants received an ASD diagnosis at 24 months of age. Due to the small sample size of infants diagnosed with ASD, all HL infants were treated as a uniform group.

**Table 1.** Sample demographics and behavioral scores.

| | HL (mean ± SD) | LL (mean ± SD) | t or $\chi^2$ |
| --- | --- | --- | --- |
| N | 62 | 39 | - |
| Sex (# female) | 22 | 15 | 0.01 (p = 0.93) |
| Scan Age (months) | 5.89 ± 0.65 | 5.97 ± 0.48 | 0.77 (p = 0.45) |
| Race <sup>a</sup> |  |  |  |
| White | 49 | 32 | 0.99 (p = 0.32) |
| Non-white or mixed-race | 11 | 3 |  |
| Ethnicity <sup>a</sup> |  |  |  |
| Hispanic | 3 | 2 | <0.001 (p = 1) <sup>b</sup> |
| Non-Hispanic | 57 | 33 |  |
| Maternal Education <sup>a</sup> |  |  |  |
| College degree & above | 42 | 31 | 3.30 (p = 0.07) |
| No college degree | 18 | 4 |  |
| Scanner head motion |  |  |  |
| Mean relative motion (mm) | 0.068 ± 0.036 | 0.07 ± 0.03 | 0.30 (p = 0.77) |
| Mean absolute motion (mm) | 0.162 ± 0.139 | 0.210 ± 0.183 | 1.40 (p = 0.17) |
| # ICA components removed | 13.2 ± 4.71 | 12.7 ± 3.59 | 0.67 (p = 0.51) |
| ISOP Score |  |  |  |
| 6 months (N) | 2.72 ± 1.22 (62) | 2.48 ± 0.793 (39) | 1.20 (p = 0.23) |
| 12 months (N) | 2.67 ± 1.04 (55) | 2.47 ± 0.937 (24) | 0.86 (p = 0.40) |
| SOR Score |  |  |  |
| 12 months (N) | 17.1 ± 5.01 (54) | 17 ± 2.67 (22) | 0.19 (p = 0.85) |
| 24 months (N) | 17.4 ± 4.70 (44) | 18.7 ± 2.31 (19) | 1.48 (p = 0.26) |
| ADOS (24-36 months) | 2.41 ± 2.31 | 1.25 ± 0.508 | 3.61 (p < 0.001) * |
| MSEL T-Scores |  |  |  |
| ELC | 93.8 ± 17.5 | 110 ± 15.5 | 4.80 (p < 0.001) * |
| Gross Motor | 46 ± 12.4 | 51.7 ± 11.7 | 2.37 (p = 0.02) * |
| Fine Motor | 48.1 ± 9.56 | 55.4 ± 9.65 | 3.73 (p < 0.001) * |
| Visual Reception | 49.8 ± 11.2 | 55.3 ± 10.6 | 2.47 (p = 0.02) * |
| Receptive Language | 45.1 ± 14 | 55.5 ± 8.58 | 4.65 (p < 0.001) * |
| Expressive Language | 43.4 ± 11.4 | 53.3 ± 10.6 | 4.45 (p < 0.001) * |

Higher Infant Sleep-Onset Problems (ISOP) scores indicate worse problems. Higher Sensory Over-
Responsivity (SOR) scores indicate higher severity SOR behavior.
\* Signifies statistical significance.
<sup>a</sup> Unknown or unreported race, ethnicity, and maternal education information for 2 HL and 3 LL infants.
<sup>b</sup> Fishers Exact Test used to confirm statistical non-significance.
ICA: Independent Component Analysis; ADOS: Autism Diagnostic Observation Schedule; MSEL: Mullen
Scales of Early Learning; ELC: Early Learning Composite

### Behavioral measures and assessments

For the present study, we used several measures related to cognitive development, adaptive functioning, and behaviors associated with ASD which were among the battery of questionnaires and assessments collected as part of the IBIS. The Infant Behavior Questionnaire Revised (IBQ-R), collected at 6 and 12 months of age, assessed temperament along 14 scales [84]. Since the original IBIS dataset did not include a sleep-specific questionnaire or assessment, five questions related to sleep-onset and settling down from the IBQ-R were summed to generate the Infant Sleep-Onset Problems (ISOP) score (Supplementary Table S1), as done in a prior study [14]. The Sensory Experiences Questionnaire (SEQ), a parent-reported measure of a child’s sensory processing patterns, was collected at 12 and 24 months [85]. A sensory over-responsivity (SOR) score was generated based on summed raw scores for auditory, visual, and tactile items that related to SOR symptoms (Supplementary Table S2). While the SEQ has a hyper-responsiveness scale, it also includes scores from one Gustatory/Olfactory question, one Vestibular/Proprioceptive question, and one addendum item on “picky” eating. Given that auditory [22], visual [86], and tactile [23] domains have reported associations with sleep problems in ASD, we restricted our SOR score to these domains. The Mullen Scales of Early Learning, collected at 6, 12, and 24 months, assessed language, motor, cognitive and perceptual abilities [83]. For most infants, age-normed scores from the 24-month assessment (N=85) were used. For infants without 24-month scores, scores from 6-month (N=7) or 12-month (N=9) assessments were used. The Autism Diagnostic Observation Schedule (ADOS), collected at 24 or 36 months was used to evaluate ASD symptoms [81].

### MRI data acquisition

MRI scans were collected during natural sleep at 6 months of age on cross-site calibrated 3T Siemens Tim Trio scanners with 12-channel head coils. In this study, we analyzed 3D T2 TSE (TR = 3,200 ms, TE = 497 ms, 160 sagittal slices, FOV = 256 mm, voxel size = 1 mm^3^) and resting-state scans (5.4 minutes each) acquired using a gradient echo planar sequence (TR =2,500 ms, TE = 27 ms, FOV = 256 mm, voxel size = 4 mm^3^, flip angle = 90) and. One to four resting-state scans were collected for each participant.

### fMRI data preprocessing

Functional MRI data were preprocessed and analyzed using FMRIB’s Software Library (FSL version 6.00; [87]). Structural T2 images were skull stripped using FSL’s Brain Extraction Tool, followed by manual inspection. Functional scans underwent motion correction and linear registration to the subject’s corresponding structural image, followed by registration to a standard 6-month infant brain template [88]. Spatial smoothing was performed using a 6-mm Gaussian kernel and then scans underwent 4D mean intensity normalization. Mean relative and absolute motion (mm) did not significantly differ between the two groups (Table 1).

Independent Component Analysis – Automatic Removal of Motion Artifacts (ICA-AROMA; [89], [90] was used to automatically remove components identified as noise or motion, as this has been shown to improve reproducibility of resting-state analyses [91]. HL and LL groups did not significantly differ on the number of ICA-AROMA components removed (Table 1). Furthermore, data were bandpass filtered (0.01 Hz < t < 0.1 Hz) to reduce physiological noise. Mean white matter, cerebrospinal fluid, and global time series were regressed from the data at the single-subject level [92].

### fMRI data analysis

All connectivity analyses were anchored at the right anterior insula (rAI) to interrogate SN connectivity, as done in several prior studies [37], [42], [44], [93], including a recent study in a similar population of infants at varying familial likelihood for ASD [44]. A rAI mask was derived from an anatomical parcellation of the right insula available from a standard infant atlas [88] . We created a rAI anatomical mask composed of voxels anterior to the midline of the center of gravity of the right insula, as done in a prior study [44]. The time series, averaged across the rAI, was extracted from processed residuals in standard space and correlated with that of every other voxel in the brain to generate single-subject functional connectivity maps. Using Fisher’s r-to-z transformation, the resulting correlation maps were converted to z-statistic maps. Group-level analyses were performed in FSL using FMRIB’s Local Analysis of Mixed Effects (FLAME 1+2), with scanner and Mullen ELC scores included as demeaned nuisance regressors. Between-group contrasts and bottom-up behavioral regressions (thresholded at Z > 2.3, and cluster corrected for multiple comparisons at P < 0.05) were masked by each anatomical region of interest (ROI), thus allowing us to specifically focus on SN connectivity with areas previously implicated in sleep disruption and sensory challenges, or shown to be altered in HL infants in prior work ([42], [44], [67], [69]. Our ROIs included cortical areas involved in sensorimotor processing (left and right sensorimotor cortex), cerebellar regions canonically known for their role in sensorimotor processing (bilateral lobules I-IV, bilateral lobules V-VI, and bilateral lobules VIIIA/B; [94], [95], [96], and cerebellar regions involved in higher order functions (left and right crus I, [74], [75] (Supplementary Fig. S1). Lobules V-VI were distinguished from lobules I-IV due to reports of their distinct functional connectivity with visual and auditory cortices [94], [97], as well as from lobules VIIIA and VIIIB due to their anatomical distance, as done in prior investigations of the sensorimotor cerebellum [67]. SN connectivity the left and right crus I was examined separately, given prior evidence implicating the right crus I in language processing and working memory [74], [75]. Sensorimotor parcellations were derived from the Shi et al. (2011) infant atlas [98]. Cerebellum parcellations were derived from an anatomical map of the adult cerebellum [99]. All parcellations were transformed to standard space before conducting single-subject analyses [88].

## Results

### Behavioral Measures

First, we examined whether HL and LL infants differed on infant sleep-onset problems (ISOP), sensory over-responsivity (SOR), and other measures of early behavioral development. There were no significant group differences in ISOP scores at 6 or 12 months (Table 1). However, HL infants showed significantly greater variance in ISOP scores relative to LL infants at 6 months (Levene’s test, p = 0.027). There were no significant group differences in SOR scores at 12 or 24 months (Table 1), nor in variance. Groups significantly differed on ASD symptomatology as assessed by the ADOS at 24-36 months (t = 3.61, p < 0.001); HL infants had higher scores, and thus greater severity of ASD-related behaviors, compared to LL infants (Fig. 1A). The groups differed on the Mullen Early Learning Composite (ELC) scores, as well as across the other Mullen subscales, such that LL infants had higher scores than HL infants (all t > 2.47, p < 0.02) (Fig. 1B). Given the significant group difference in Mullen ELC scores, these were added as a regressor in all analyses to ensure that any significant findings were not related to Mullen ELC differences.

**Fig 1.**
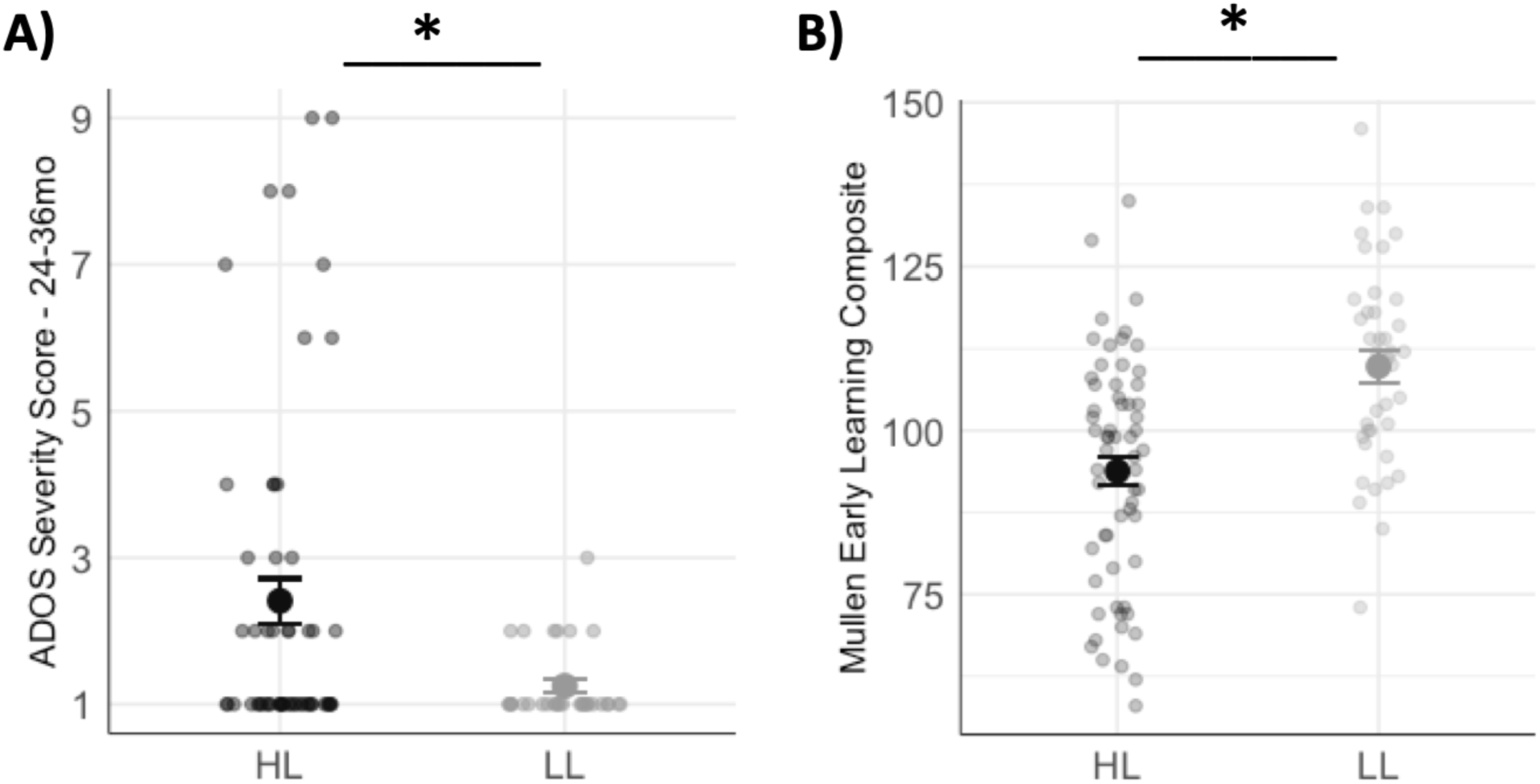
Differences in developmental scores between HL and LL infants. **A)** Significant group difference in Autism Diagnostic Observation Schedule (ADOS) severity scores from 24-36 months (p < 0.001). **B)** Significant group difference in Mullen early language composite scores (p < 0.001).

Next, we investigated relationships between sleep-onset problems and sensory over-responsivity, as well as their associations with later ASD symptomatology. There was a significant concurrent relationship between ISOP and SOR scores at 12 months in HL infants but not LL infants, such that greater sleep-onset problems were related to higher sensory over-responsivity in HL infants only (Spearman’s *ρ* = 0.34, p = 0.013) (Fig. 2A). However, there was no significant association between 6- or 12-month ISOP scores and later SOR scores in either group (Table 2). Finally, we asked how ISOP and SOR related to later ASD symptomatology. In HL infants, greater ISOP scores at 6 months (Spearman’s *ρ* = 0.27, p = 0.045) and greater SOR scores at 12 months (Spearman’s *ρ* = 0.35, p = 0.015) predicted greater ADOS severity scores at 24–36-month outcome (Fig. 2B, C). LL infants did not display significant correlations between ASD symptomatology and sleep-onset problems or sensory over-responsivity (Table 2).

**Fig 2.**
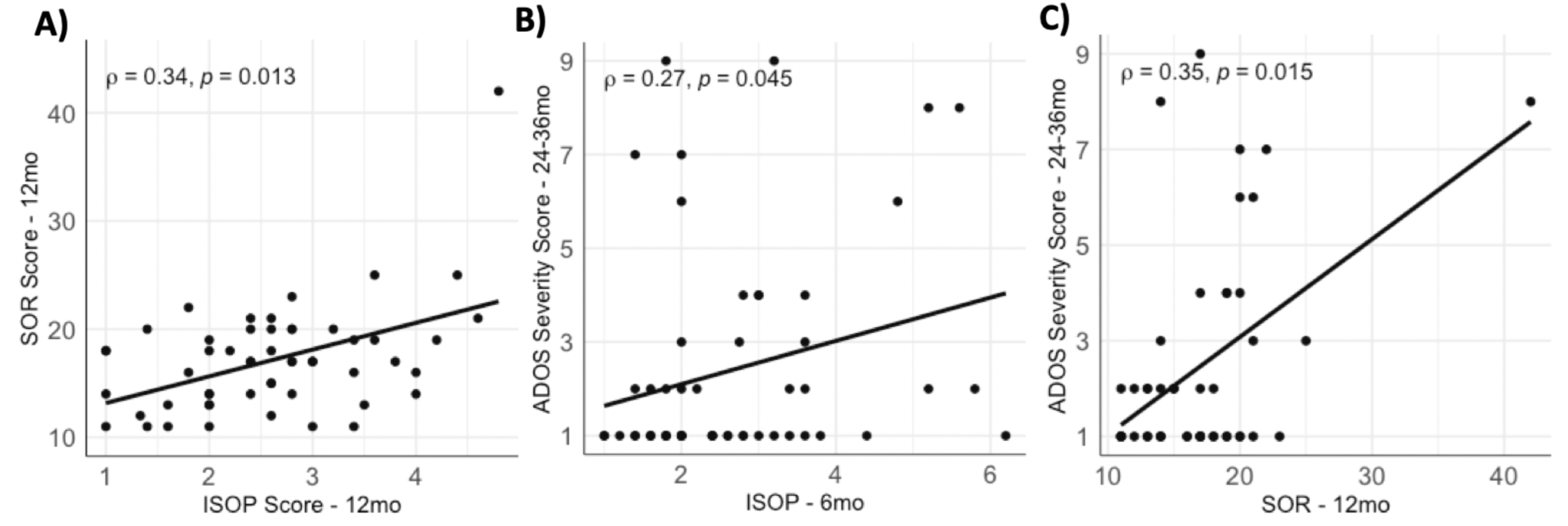
Spearman rank order correlations between Infant Sleep-Onset Problems (ISOP), Sensory Over-responsivity (SOR), and Autism Diagnostic Observation Schedule (ADOS) Scores in HL infants. **A)** Significant correlation between ISOP and SOR at 12 months (p = 0.013). **B)** Significant correlation between 6-month ISOP and ADOS severity scores at 24-36 months (p = 0.045). **C)** Significant correlation between 12-month SOR and ADOS severity scores at 24-36 months (p = 0.015).

**Table 2.**
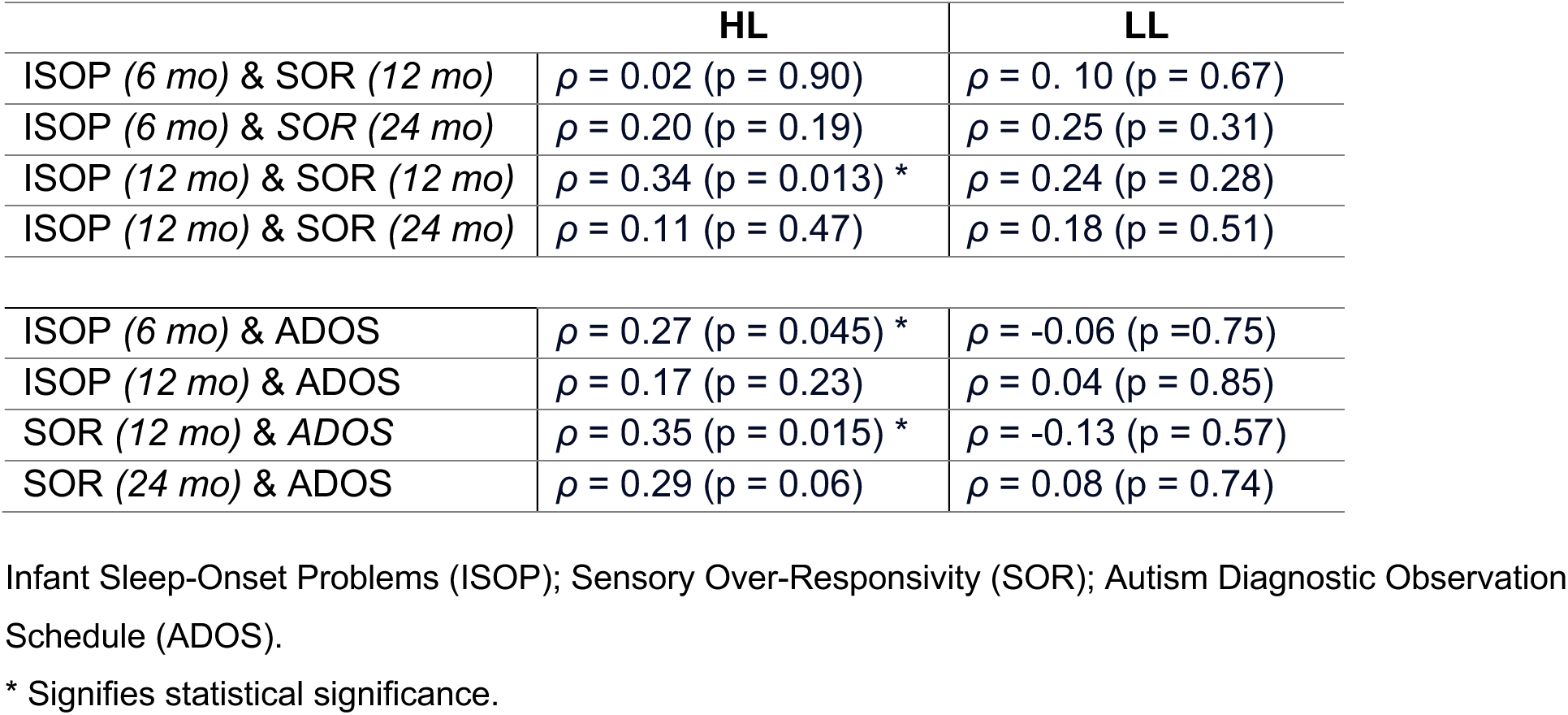
Spearman rank order correlations between ISOP, SOR, and ADOS scores.

### Group differences in salience network connectivity

Prior work has shown altered SN connectivity with sensorimotor regions in HL infants [44] and altered cerebellar connectivity in infants with delayed language trajectories, many of whom had a family history of ASD [69]. Therefore, we used an *a priori* approach and focused our analyses on these regions of interest: left and right sensorimotor cortex, bilateral cerebellum lobules I-IV, bilateral cerebellum lobules V-VI, bilateral cerebellum lobules VIIIA/B, left and right cerebellum crus I. Between-group comparisons revealed that LL infants showed stronger SN connectivity with the left cerebellum crus I and left lobules V-VI in comparison to HL infants, indicating weaker connectivity between the SN and cerebellar regions involved in sensorimotor processing and higher-order cognition in infants at elevated liability for ASD. In addition, HL infants showed stronger SN connectivity with the left sensorimotor cortex in comparison to LL infants (Fig. 3; see Table 3 for coordinate table).

**Fig 3.**
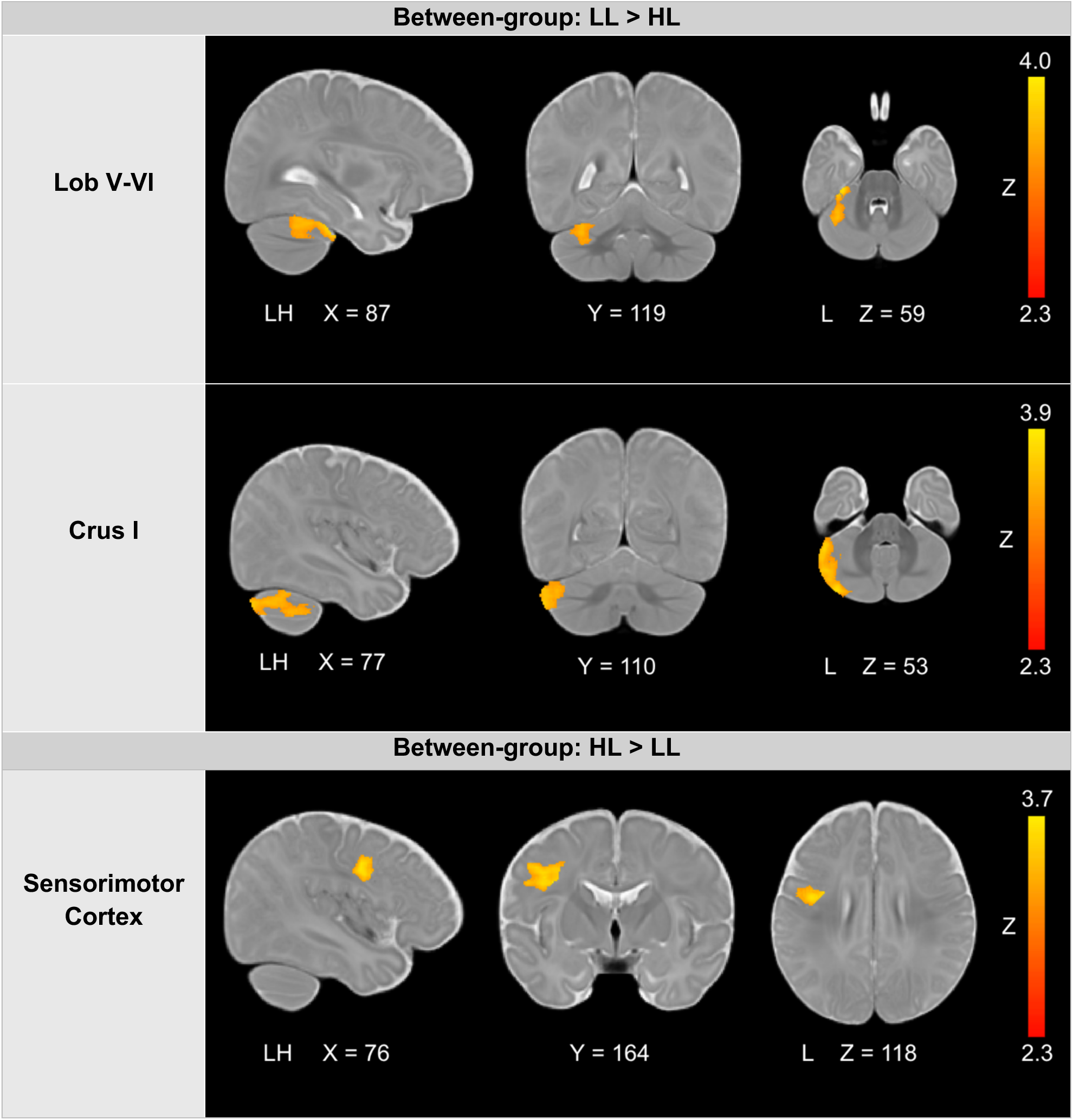
Between-group comparisons of SN connectivity to cerebellar regions lobules V-VI and crus I, as well as sensorimotor cortex. Results were thresholded at Z >2.3 and cluster-corrected for multiple comparisons at p < 0.05.

**Table 3.** Coordinate table for functional connectivity peaks in group comparisons and behavioral regressions.

|  | Cluster location | L/R | Max Z | Peak (voxel coordinates) |  |  |
| --- | --- | --- | --- | --- | --- | --- |
|  |  |  |  | x | y | z |
| LL > HL | Crus I | L | 3.94 | -40.7 | -31.6 | -32.7 |
| LL > HL | Lobule V-VI | L | 4.07 | -24.7 | -19.6 | -28.7 |
| HL > LL | Sensorimotor cortex | L | 3.72 | -33.5 | -4.4 | 25.8 |
| HL, ISOP + at 6 mo | Sensorimotor cortex | R | 3.02 | 18.6 | -19.6 | 53.9 |
| HL, ISOP+ at 12 mo | Lobule VIIIA/B | L | 2.94 | -12.6 | -46 | -39.1 |
| HL, SOR + at 12 mo | Sensorimotor cortex | L | 3.9 | -35.1 | -22 | 47.5 |
| HL, SOR + at 12 mo | Crus I | R | 3.13 | 27.5 | -46 | -27.1 |
| LL, ISOP – at 6 mo | Crus I | R | 3.85 | 28.3 | -53.2 | -34.3 |
| LL, SOR – at 12 mo | Lobule I-IV | L | 3.52 | -15.8 | -24.4 | -22.3 |
| HL>LL, SOR at 12mo | Lobule V-VI | L | 4.91 | -18.2 | -24.4 | -23.9 |
| HL, SOR + at 12 mo | Lobule V-VI | L | 3.55 | -16.6 | -40.4 | -25.5 |
| LL, SOR – at 12mo | Lobule V-VI | L | 4.78 | -19 | -25.2 | -23.9 |

### Sleep-onset problem associations with salience network connectivity

To determine whether SN connectivity with our regions of interest was associated with sleep-onset problems, ISOP scores (collected at 6 and 12 months) were entered as regressors in group connectivity analyses. In HL infants, stronger SN connectivity with right sensorimotor cortex was associated with worse concurrent sleep-onset problems at 6 months (Fig. 4A). These results were present bilaterally at lower, uncorrected thresholds. Furthermore, in HL infants, there was a positive association between SN connectivity with left lobules VIIIA/B and ISOP scores at 12 months, such that stronger SN-lobules VIIIA/B connectivity predicted worse sleep-onset problems (Fig. 4C). In LL infants, there was a negative concurrent association between SN connectivity with right crus I and ISOP scores at 6 months in LL infants, such that weaker SN-crus I connectivity was associated with worse concurrent sleep-onset problems (Fig. 4E). These findings indicate that stronger SN connectivity to sensory processing regions was associated with worse concurrent sleep-onset problems and that stronger SN connectivity to cerebellar sensory learning regions predicted later sleep-onset problems in HL infants. In contrast, stronger SN connectivity to higher-order cerebellar regions was related to fewer concurrent sleep-onset problems in LL infants.

**Fig 4.**
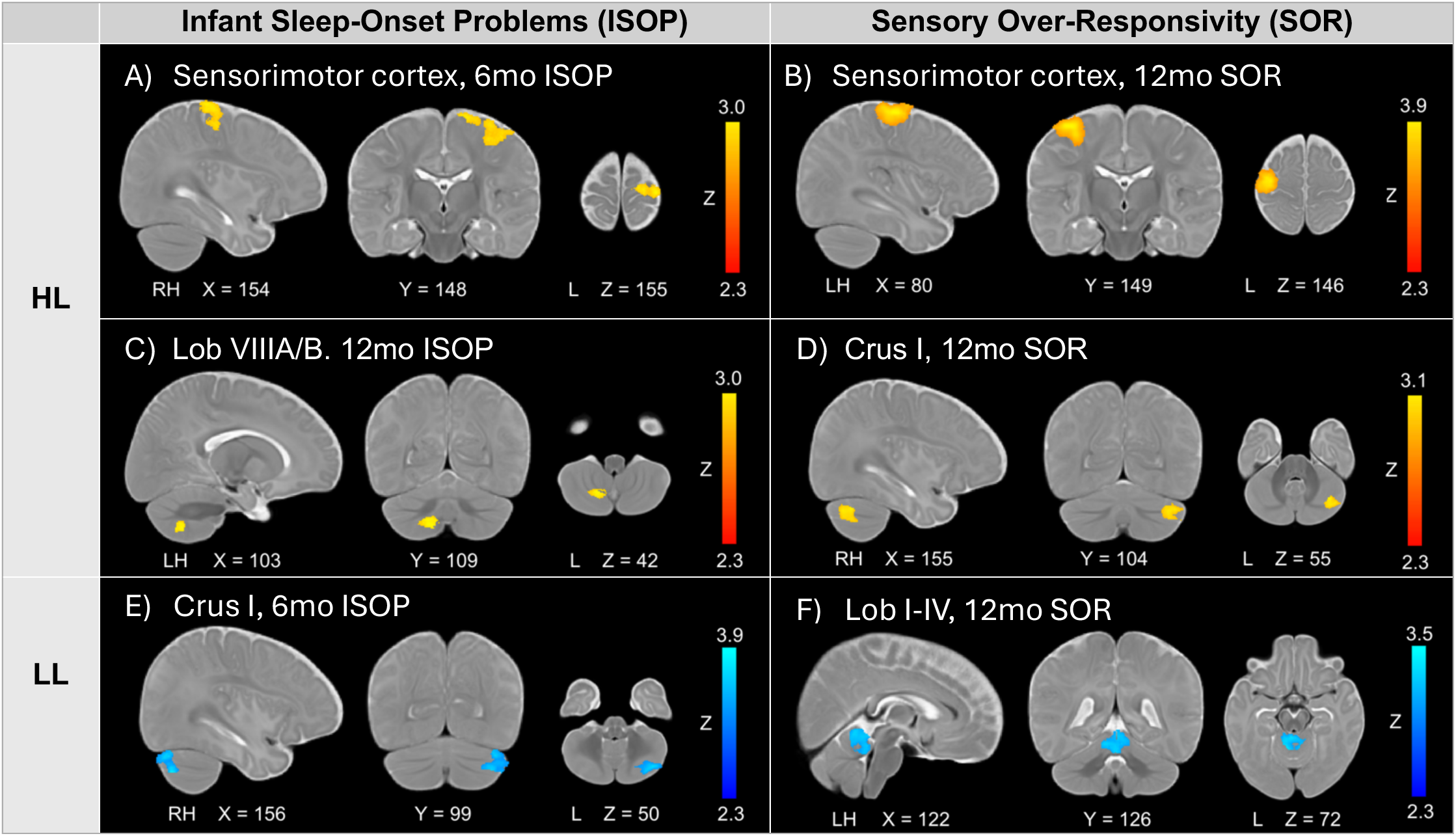
SN connectivity with *a priori* regions of interest as a function of Infant Sleep-Onset Problems (ISOP) at 6 and 12 months, and Sensory Over-Responsivity (SOR) at 12 months. Warm color scale indicates positive relationships between connectivity and behavioral scores (i.e., stronger connectivity associated with worse ISOP or SOR). Cool color scale indicates negative relationships between connectivity and behavioral scores (i.e., weaker connectivity associated with worse ISOP or SOR). Contrasts were thresholded at Z > 2.3 and cluster-corrected for multiple comparisons at p < 0.05.

### Sensory over-responsivity associations with salience network connectivity

SOR scores (at 12 and 24 months) were used as regressors to examine associations with SN connectivity. In HL infants, stronger SN connectivity with left sensorimotor cortex predicted greater SOR severity at 12 months of age (Fig. 4B). These results were present bilaterally at lower, uncorrected thresholds. Additionally, stronger SN connectivity with right crus I predicted greater SOR severity at 12 months in HL infants (Fig. 4D). In LL infants, weaker SN connectivity with lobules I-IV predicted worse SOR severity at 12 months (Fig. 4F). Finally, there was a significant group interaction between SN connectivity with lobules V-VI and 12-month SOR (Fig. 5). This interaction reflected that HL infants had a positive association between SN connectivity with lobules V-VI and 12-month SOR, with stronger connectivity predicting worse SOR severity, whereas LL infants had a negative association between SN connectivity with lobules V-VI and 12-month SOR, with weaker connectivity predicted worse SOR severity. No associations were observed with 24-month SOR. Overall, stronger SN connectivity to sensory processing cortices and cerebellar regions involved in sensory prediction and higher-order functions predicted later SOR in HL infants. In contrast, stronger SN connectivity to the sensory learning cerebellum predicted lower SOR at toddlerhood in LL infants.

**Fig 5.**
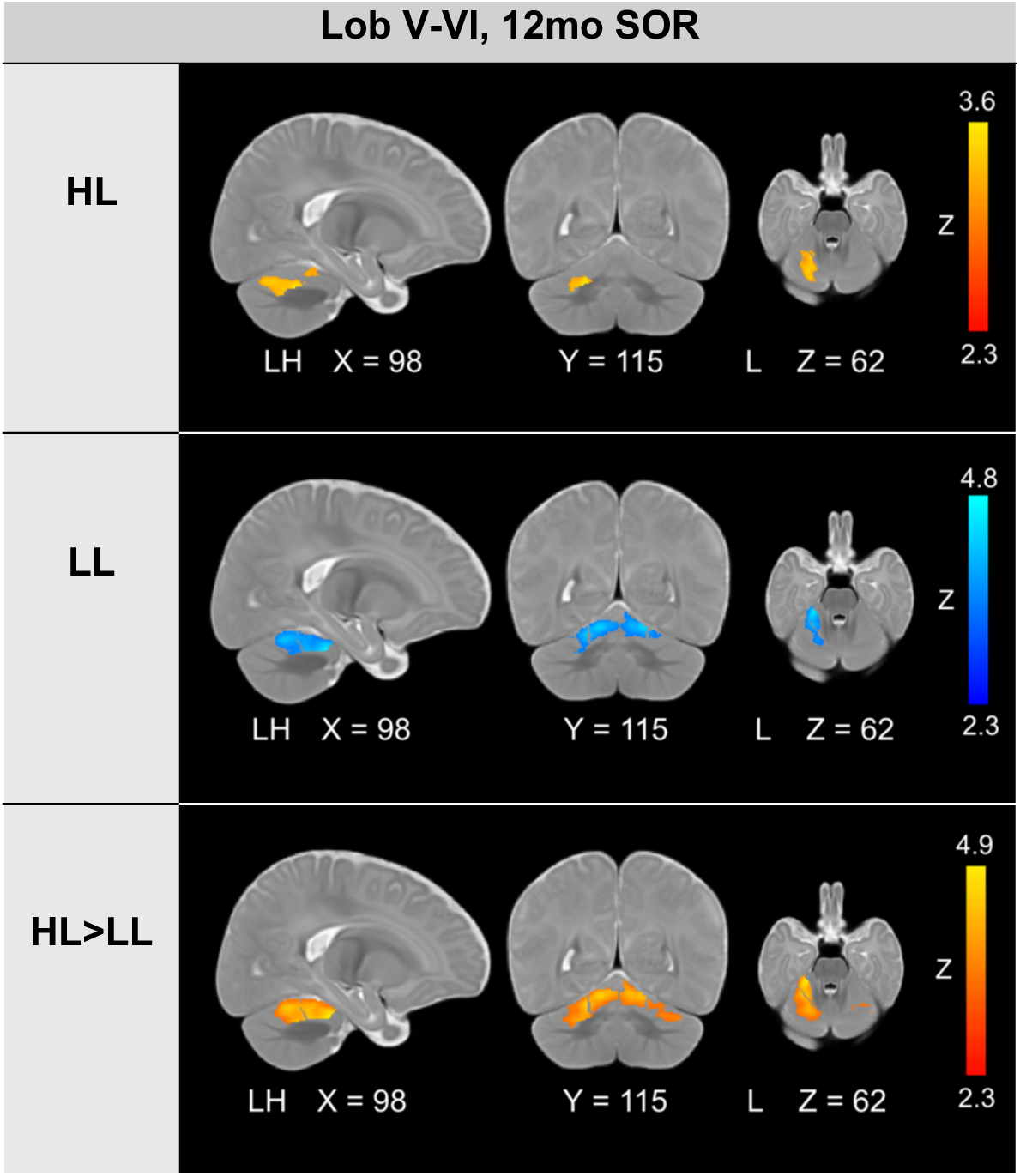
Group interaction of SN connectivity with cerebellar lobules V-VI and Sensory Over-Responsivity (SOR) at 12 months. Warm color scale indicates positive relationships between connectivity and behavioral scores (i.e., stronger connectivity associated with worse ISOP or SOR). Cool color scale indicates negative relationships between connectivity and behavioral scores (i.e., weaker connectivity associated with worse ISOP or SOR). Contrasts were thresholded at Z > 2.3 and cluster-corrected for multiple comparisons at p < 0.05.

## Discussion

In the present study, we investigated functional connectivity between a hub of the salience network (SN) and sensorimotor cortices, as well as with the sensorimotor and higher-order cerebellum, as a possible mechanism underlying sleep-onset problems and sensory over-responsivity (SOR) in 6-month-old infants at high familial likelihood for ASD. Overall, HL and LL infants differed in their SN connectivity with sensorimotor cortex and cerebellar lobules V-VI and crus I. We also examined how SN connectivity patterns related to sleep-onset problems and SOR. LL infants with milder sleep-onset problems and lower SOR demonstrated stronger SN connectivity to cerebellar sensorimotor regions (lobules I-IV, V-VI) and crus I. Interestingly, HL infants displayed an opposite pattern of associations whereby infants with worse sleep-onset problems and greater SOR had stronger SN connectivity with sensorimotor regions (sensorimotor cortex, cerebellar lobules V-VI, VIIIA/B) and cerebellum crus I.

We first examined sleep-onset patterns, sensory processing, and ASD symptomatology in HL and LL infants to understand how their developmental trajectories might diverge. Consistent with our predictions, HL infants displayed a significant positive association between sleep-onset problems and SOR at 12 months, such that worse sleep-onset problems were correlated with more severe SOR. This is in line with findings in older ASD youth and adults [26], [86], [100], and previous reports in children as young as toddler and preschool age [22], [25], [27]. Our findings indicate that co-occurrence of sleep problems and sensory challenges manifests earlier in development than previously shown. Furthermore, worse sleep onset problems at 6 months and higher SOR at 12 months both predicted greater subsequent ASD symptomatology at 24-36 months in HL infants, suggesting that altered sleep and sensory processing in HL infants is predictive of ASD symptoms 1-1.5 years later. These results support retrospective reports of early sleep problems that present prior to overt ASD symptoms [2] and converge with previous literature showing a relationship between early sensory challenges and later ASD traits [20], [101], [102], [103]. Although HL and LL infants did not differ in overall sleep-onset challenges or SOR, the HL group exhibited greater variability in sleep-onset problems, reflecting increased behavioral heterogeneity. This variability aligns with the diverse outcomes observed in HL infants – while some HL infants will later receive an ASD diagnosis, others may show developmental delays [47], [104], subclinical traits, or typical development. The lack of group differences, or variability, in SOR might be attributed to challenges that parents may face in reporting a child’s SOR in early life, prior to language acquisition which would allow a child to verbalize discomfort for certain sensory stimuli. Despite comparable mean levels of sleep and sensory challenges, the groups significantly differed on ASD symptomatology, as assessed by the ADOS, and broader developmental measures (e.g., motor, language, visual processing), as assessed by the Mullen. These developmental differences, as well as the unique relationship observed between sleep-onset problems and SOR in HL infants, underscore the need to identify early pathways of brain development that shape divergent developmental trajectories.

Given the well-established relationship between sleep disruption and sensory challenges in ASD [22], [25], [26], [27], [86], [100], our functional connectivity analyses focused on SN connectivity with the sensorimotor cortex as a potential neural mechanism underlying this association. In comparison to LL controls, HL infants displayed stronger SN connectivity with sensorimotor cortex. Interestingly, this stronger SN connectivity with sensorimotor cortices at 6 months related to worse concurrent sleep-onset problems and predicted greater SOR at 12 months in HL infants. At a mechanistic level, this atypical pattern of SN connectivity could result in increased attribution of salience to sensory stimuli in HL infants, which may not only lead to sensory processing challenges, but also disrupt sleep. Our findings directly corroborate recent work [44], which has shown that as early as 6 weeks old, HL infants already exhibit stronger SN connectivity with regions involved in sensorimotor processing, and that stronger SN connectivity with primary auditory and sensory processing regions predict greater SOR in HL infants. Notably, we significantly expand on this prior work by showing that this pattern of SN hyperconnectivity with sensorimotor cortex persists at 6 months of age and is also associated with sleep-onset problems in HL infants. These findings are also in line with prior reports of stronger SN connectivity with sensorimotor cortex in adult patients with insomnia [76, p. 202] and in ASD youth with SOR [42], [43], suggesting that this pattern may linger across the lifespan and may also relate to sleep disorders more broadly. This shared aspect of atypical SN connectivity may contribute to the distinct “hyperarousal” theories posited in both the ASD and insomnia literatures, whereby increased arousal to external or internal stimuli may contribute to both core ASD symptoms [105] and sleep difficulties [106], respectively. Yet, this pattern of SN hyperconnectivity in older individuals has only been investigated in relation to sleep disruption and sensory challenges independently. To our knowledge, this study is the first to directly implicate SN-sensorimotor cortex connectivity as a shared neural mechanism underlying both sleep disturbances and sensory processing atypicalities, suggesting that a possible convergent relationship may already be present very early in development.

Recent work implicating altered sensorimotor cerebellar connectivity in ASD youth with SOR [67] and atypical crus I connectivity patterns in an infant sample with a family history of ASD [69] prompted us to also explore differences in SN connectivity with these cerebellar regions. Overall, in comparison to LL controls, HL infants exhibited weaker SN connectivity with the sensorimotor cerebellum (lobules V-VI), as well as with crus I. Interestingly, lobule VI and crus I have been shown to be intrinsically connected to the SN in neurotypical adults [107]. However, autistic adults show reduced connectivity between the rostral prefrontal cortex, another cortical node of the SN, and cerebellar lobule VI and crus I [40]. Our findings show that this pattern of connectivity is already present in LL infants and disrupted in HL infants, suggesting that aberrant SN-cerebellar connectivity is present as early as 6 months of age in infants at increased likelihood for ASD. Furthermore, our findings indicate that in LL infants, stronger SN connectivity with cerebellar areas at 6 months predicts more favorable sensory and sleep outcomes. Specifically, stronger connectivity between the SN and the sensorimotor cerebellum (lobules I-IV and lobules V-VI) was associated with less sensory over-responsivity at 12 months, while stronger connectivity with right crus I was associated with fewer concurrent sleep-onset problems. Given the cerebellum’s important role in error-based and predictive learning [108], increased functional coupling with the SN may enable early sensory learning which, in turn, may lead to more developmentally appropriate processing of external stimuli. Interestingly, in HL infants, stronger SN connectivity with some unique and overlapping cerebellar regions predicted worse scores on 12-month measures of sensory over-responsivity and sleep-onset problems. Here, stronger SN connectivity with lobules V-VI and the right crus I predicted worse later sensory over-responsivity, whereas stronger SN connectivity with lobules VIIIA/B predicted worse later sleep-onset problems in HL infants. Cerebellar dysfunction at the structural and functional level has consistently been reported in ASD [62], [67], [109], [110], [111], [112], [113], potentially triggering atypicalities in error prediction and learning. Canonically known for its role in motor control, the cerebellum utilizes error signals produced through mismatches between true and predicted outcomes to refine its internal model for future predictions [95], [114]. However, increasing evidence shows that the cerebellum’s role in prediction learning applies to other domains beyond motor control [115], [116], [117]. Taken together, these results suggest that in HL infants, insufficient predictive learning may hinder sensory habituation and cause heightened reactions to external stimuli. Thus, strengthening of SN-cerebellar networks, while conducive to sensory learning in typical development, may result in sustained and heightened salience of external stimuli which are not properly integrated into an updated prediction model in HL infants, ultimately contributing to SOR and sleep problems.

Overall, these findings suggest that HL infants show atypicalities in the functional networks involved in the detection and filtering of sensory stimuli, and related predictive learning. Given that proper sensory gating is responsible for facilitating and maintaining sleep [34], improper allocation of attention to irrelevant sensory inputs and perturbed sensory prediction may interfere with the ability to fall asleep. Indeed, recent work has implicated functional overconnectivity of sensory cortices with the thalamus – the brain’s sensory relay station – in sensory sensitivities in HL infants [59] and sleep problems in autistic toddlers [22]. Here, we identify the SN as a key intrinsic functional network linking early sleep problems and SOR, and add to a growing body of work showing early functional connectivity atypicalities in infants at high familial likelihood for ASD [44], [45], [46], [50], [51], [52], [56], [57], [58], [59], [118]. We show that HL infants exhibit an altered pattern of SN connectivity that persists at 6 months of age, which may predispose them to directing undue attention to sensory stimuli, thus interfering with sensory prediction and leading to sleep and sensory processing challenges. This emphasizes the need for early intervention to alleviate these emerging atypicalities and their potential downstream behavioral effects. While the etiology of sleep problems in ASD is likely multifactorial (e.g., circadian rhythm disruption, behavioral dysregulation) [119], our findings suggest that limiting sensory stimuli during bedtime could be one possible intervention. For instance, sensory modifications, such as blackout curtains to reduce visual stimuli or earplugs to attenuate auditory stimuli, may help improve sleep onset and sleep maintenance. As proper sleep is crucial for brain development [120], implementing early interventions while the brain is still highly plastic may have dramatic effects on later outcomes [121], [122] and scaffold optimal neurodevelopment.

### Limitations

This study has several limitations. First, given the small number of infants who received an ASD diagnosis (N = 7), our analyses were focused on the HL group as a whole. Accordingly, our findings offer new insights into altered developmental trajectories in infants at increased familial likelihood for ASD, as opposed to defining differences based on later diagnostic outcome. Second, our investigation of the SN was limited to only one of its main hubs, the right anterior insula, consistent with previous studies of the SN [37], [42], [44], [93]. Future work examining other SN nodes (e.g. anterior cingulate) may provide a more comprehensive understanding of SN connectivity. Third, while parent-reported questionnaires are generally reliable [123] and are the most common method used to assess sleep patterns and sensory processing in infants and young children, the lack of more objective measures presents another limitation. Future research incorporating objective tools, such as actigraphy for sleep [124] and clinician-administered assessments for sensory processing (e.g., Sensory Assessment for Neurodevelopmental Disorders [SAND]; [125] could provide critical insights into the early development of sleep and sensory challenges. Fourth, while sensory processing atypicalities in ASD encompass SOR, sensory hypo-responsivity and sensory seeking behaviors [126], we focused only on SOR given its previously reported associations with sleep problems [23], [24], [86]. Lastly, as in most infant neuroimaging studies to date, the infants’ sleep stage was not assessed during the MRI scan, primarily due to the considerable challenge of collecting simultaneous EEG and MRI data in this population [127]. However, studies collecting MRI data during natural sleep in infants and toddlers take several steps to minimize this possible confound (e.g., Linke et al., 2023 [22]).

## Conclusions

In summary, by leveraging functional MRI in infants at high and low likelihood for ASD, these findings advance our understanding of how diverging patterns of early brain functional connectivity patterns may contribute to altered neurodevelopmental trajectories, ultimately driving the sleep and sensory challenges widely observed in ASD. We found that HL infants exhibited atypical SN connectivity to sensorimotor cortices and cerebellar regions that was associated with more severe sleep-onset problems and sensory over-responsivity. These sleep and sensory processing challenges in HL infants – which were behaviorally linked to one another and to later ASD symptomatology – may share a common neural substrate as early as 6 months of age. This work expands on prior reports of altered SN connectivity associated with ASD and sleep disruption in older individuals and suggests that over-attribution of attention to sensory stimuli and aberrant prediction learning manifest in early infancy. Our findings suggest that reducing sensory stimuli may be one solution to mitigate sensory over-responsivity and sleep disruption during a period of heightened neuroplasticity, when adequate sleep may exert a particularly strong influence on subsequent neurodevelopment.

## Acknowledgements

We are grateful for the families and children who participated in the Infant Brain Imaging Study (IBIS). Thank you to Dr. John Pruett and the IBIS steering committee for their support on this project.

## Data Availability

The data used in these analyses are available via the National Institutes of Mental Health Data Archive (collections #19 and #2027). Researchers can submit a Data Access Request to access de-identified human subjects data at https://nda.nih.gov/.

## Authors’ contributions

EC: methodology, data processing, data analysis, visualization, writing original draft, writing – review and editing, revision and finalization; LW: writing – review and editing, revision and finalization; LMH: writing – review and editing, revision and finalization; SG: writing – review and editing, revision and finalization; MD: funding acquisition, conceptualization, supervision, writing – review and editing.

## References

[1] M. J. Maenner et al., “Prevalence and Characteristics of Autism Spectrum Disorder Among Children Aged 8 Years - Autism and Developmental Disabilities Monitoring Network, 11 Sites, United States, 2018,” Morb. Mortal. Wkly. Rep. Surveill. Summ. Wash. DC 2002, vol. 70, no. 11, pp. 1–16, Dec. 2021, doi: 10.15585/mmwr.ss7011a1.

[2] V. Guinchat et al., “Very early signs of autism reported by parents include many concerns not specific to autism criteria,” Res. Autism Spectr. Disord., vol. 6, no. 2, pp. 589–601, Apr. 2012, doi: 10.1016/j.rasd.2011.10.005.

[3] American Psychiatric Association. Diagnostic and statistical manual of mental disorders (5th ed.). 2013.

[4] S. Cohen, R. Conduit, S. W. Lockley, S. M. Rajaratnam, and K. M. Cornish, “The relationship between sleep and behavior in autism spectrum disorder (ASD): a review,” J. Neurodev. Disord., vol. 6, no. 1, p. 44, 2014, doi: 10.1186/1866-1955-6-44.

[5] A. Shaw, T. N. T. Do, L. Harrison, M. Marczak, D. Dimitriou, and A. Joyce, “Sleep and Cognition in People with Autism Spectrum Condition: A Systematic Literature Review,” Rev. J. Autism Dev. Disord., vol. 9, no. 3, pp. 416–426, Sep. 2022, doi: 10.1007/s40489-021-00266-7.

[6] M. Pisch, F. Wiesemann, and A. Karmiloff-Smith, “Infant wake after sleep onset serves as a marker for different trajectories in cognitive development,” J. Child Psychol. Psychiatry, vol. 60, no. 2, pp. 189–198, 2019, doi: 10.1111/jcpp.12948.

[7] W. Sun et al., “A Community-Based Study of Sleep and Cognitive Development in Infants and Toddlers,” J. Clin. Sleep Med. JCSM Off. Publ. Am. Acad. Sleep Med., vol. 14, no. 6, pp. 977–984, Jun. 2018, doi: 10.5664/jcsm.7164.

[8] G. M. Mason, S. Lokhandwala, T. Riggins, and R. M. C. Spencer, “Sleep and human cognitive development,” Sleep Med. Rev., vol. 57, p. 101472, Jun. 2021, doi: 10.1016/j.smrv.2021.101472.

[9] E. Dearing, K. McCartney, N. L. Marshall, and R. M. Warner, “Parental reports of children’s sleep and wakefulness: longitudinal associations with cognitive and language outcomes,” Infant Behav. Dev., vol. 24, no. 2, pp. 151–170, Feb. 2001, doi: 10.1016/S0163-6383(01)00074-1.

[10] K. Horváth and K. Plunkett, “Frequent daytime naps predict vocabulary growth in early childhood,” J. Child Psychol. Psychiatry, vol. 57, no. 9, pp. 1008–1017, Sep. 2016, doi: 10.1111/jcpp.12583.

[11] X. Liang, X. Zhang, Y. Wang, M. H. van IJzendoorn, and Z. Wang, “Sleep problems and infant motor and cognitive development across the first two years of life: The Beijing Longitudinal Study,” Infant Behav. Dev., vol. 66, p. 101686, Feb. 2022, doi: 10.1016/j.infbeh.2021.101686.

[12] J. Begum-Ali et al., “Infant sleep predicts trajectories of social attention and later autism traits,” J. Child Psychol. Psychiatry, vol. 64, no. 8, pp. 1200–1211, 2023, doi: 10.1111/jcpp.13791.

[13] K. Kikuchi et al., “Sleep quality and temperament in association with autism spectrum disorder among infants in Japan,” Commun. Med., vol. 3, no. 1, pp. 1–8, Jun. 2023, doi: 10.1038/s43856-023-00314-9.

[14] K. E. MacDuffie et al., “Sleep-onset problems and subcortical development in infants later diagnosed with autism spectrum disorder,” Am. J. Psychiatry, vol. 177, no. 6, pp. 518–525, Jun. 2020, doi: 10.1176/appi.ajp.2019.19060666.

[15] A. Ben-Sasson et al., “Extreme sensory modulation behaviors in toddlers with autism spectrum disorders,” Am. J. Occup. Ther. Off. Publ. Am. Occup. Ther. Assoc., vol. 61, no. 5, pp. 584–592, 2007, doi: 10.5014/ajot.61.5.584.

[16] T. B. Carson, M. J. Valente, B. J. Wilkes, and L. Richard, “Brief Report: Prevalence and Severity of Auditory Sensory Over-Responsivity in Autism as Reported by Parents and Caregivers,” J. Autism Dev. Disord., vol. 52, no. 3, pp. 1395–1402, Mar. 2022, doi: 10.1007/s10803-021-04991-0.

[17] E. Campi et al., “Sensory Reactivity of Infants at Elevated Likelihood of Autism and Associations with Caregiver Responsiveness,” J. Autism Dev. Disord., vol. 54, no. 1, pp. 270–279, Jan. 2024, doi: 10.1007/s10803-022-05764-z.

[18] S. A. Green, A. Ben-Sasson, T. W. Soto, and A. S. Carter, “Anxiety and sensory over-responsivity in toddlers with autism spectrum disorders: bidirectional effects across time,” J. Autism Dev. Disord., vol. 42, no. 6, pp. 1112–1119, Jun. 2012, doi: 10.1007/s10803-011-1361-3.

[19] A. Ben-Sasson, T. W. Soto, F. Martínez-Pedraza, and A. S. Carter, “Early sensory over-responsivity in toddlers with autism spectrum disorders as a predictor of family impairment and parenting stress,” J. Child Psychol. Psychiatry, vol. 54, no. 8, pp. 846–853, 2013, doi: 10.1111/jcpp.12035.

[20] J. J. Wolff et al., “A longitudinal study of parent-reported sensory responsiveness in toddlers at-risk for autism,” J. Child Psychol. Psychiatry, vol. 60, no. 3, pp. 314–324, Mar. 2019, doi: 10.1111/jcpp.12978.

[21] E. Worthley et al., “Sensory Profiles in Relation to Later Adaptive Functioning Among Toddlers at High-Familial Likelihood for Autism,” J. Autism Dev. Disord., vol. 54, no. 6, pp. 2183–2197, Jun. 2024, doi: 10.1007/s10803-022-05869-5.

[22] A. C. Linke et al., “Sleep Problems in Preschoolers With Autism Spectrum Disorder Are Associated With Sensory Sensitivities and Thalamocortical Overconnectivity,” Biol. Psychiatry Cogn. Neurosci. Neuroimaging, vol. 8, no. 1, pp. 21–31, Jan. 2023, doi: 10.1016/j.bpsc.2021.07.008.

[23] O. Tzischinsky et al., “Sleep disturbances are associated with specific sensory sensitivities in children with autism,” Mol. Autism, vol. 9, p. 22, Mar. 2018, doi: 10.1186/s13229-018-0206-8.

[24] M. O. Mazurek and G. F. Petroski, “Sleep problems in children with autism spectrum disorder: examining the contributions of sensory over-responsivity and anxiety,” Sleep Med., vol. 16, no. 2, pp. 270–279, Feb. 2015, doi: 10.1016/j.sleep.2014.11.006.

[25] M. O. Mazurek, K. Dovgan, A. M. Neumeyer, and B. A. Malow, “Course and Predictors of Sleep and Co-occurring Problems in Children with Autism Spectrum Disorder,” J. Autism Dev. Disord., vol. 49, no. 5, pp. 2101–2115, May 2019, doi: 10.1007/s10803-019-03894-5.

[26] S. Reynolds, S. J. Lane, and L. Thacker, “Sensory Processing, Physiological Stress, and Sleep Behaviors in Children with and without Autism Spectrum Disorders,” OTJR, vol. 32, no. 1, pp. 246–257, Jan. 2012, doi: 10.3928/15394492-20110513-02.

[27] L. Manelis-Baram et al., “Sleep Disturbances and Sensory Sensitivities Co-Vary in a Longitudinal Manner in Pre-School Children with Autism Spectrum Disorders,” J. Autism Dev. Disord., vol. 52, no. 2, pp. 923–937, 2022, doi: 10.1007/s10803-021-04973-2.

[28] M. D. Thye, H. M. Bednarz, A. J. Herringshaw, E. B. Sartin, and R. K. Kana, “The impact of atypical sensory processing on social impairments in autism spectrum disorder,” Dev. Cogn. Neurosci., vol. 29, pp. 151–167, Jan. 2018, doi: 10.1016/j.dcn.2017.04.010.

[29] J. M. Saletin et al., “Sleep Problems and Autism Impairments in a Large Community Sample of Children and Adolescents,” Child Psychiatry Hum. Dev., vol. 55, no. 5, pp. 1167–1175, Oct. 2024, doi: 10.1007/s10578-022-01470-0.

[30] S. E. Schulz and R. A. Stevenson, “Sensory hypersensitivity predicts repetitive behaviours in autistic and typically-developing children,” Autism Int. J. Res. Pract., vol. 23, no. 4, pp. 1028–1041, May 2019, doi: 10.1177/1362361318774559.

[31] Y.-Q. Kang, X.-R. Song, G.-F. Wang, Y.-Y. Su, P.-Y. Li, and X. Zhang, “Sleep Problems Influence Emotional/Behavioral Symptoms and Repetitive Behavior in Preschool-Aged Children With Autism Spectrum Disorder in the Unique Social Context of China,” Front. Psychiatry, vol. 11, p. 273, Apr. 2020, doi: 10.3389/fpsyt.2020.00273.

[32] J. A. Hollway and M. G. Aman, “Sleep correlates of pervasive developmental disorders: A review of the literature,” Res. Dev. Disabil., vol. 32, no. 5, pp. 1399–1421, Sep. 2011, doi: 10.1016/j.ridd.2011.04.001.

[33] J. A. Hollway, M. G. Aman, and E. Butter, “Correlates and risk markers for sleep disturbance in participants of the Autism Treatment Network,” J. Autism Dev. Disord., vol. 43, no. 12, pp. 2830–2843, Dec. 2013, doi: 10.1007/s10803-013-1830-y.

[34] A. Coenen, “Sensory gating and gaining in sleep: the balance between the protection of sleep and the safeness of life (a review),” J. Sleep Res., vol. 33, no. 5, p. e14152, 2024, doi: 10.1111/jsr.14152.

[35] M. C. Souders et al., “Sleep in Children with Autism Spectrum Disorder,” Curr. Psychiatry Rep., vol. 19, no. 6, p. 34, May 2017, doi: 10.1007/s11920-017-0782-x.

[36] E. Turk et al., “Functional Connectome of the Fetal Brain,” J. Neurosci., vol. 39, no. 49, pp. 9716–9724, Dec. 2019, doi: 10.1523/JNEUROSCI.2891-18.2019.

[37] W. W. Seeley et al., “Dissociable Intrinsic Connectivity Networks for Salience Processing and Executive Control,” J. Neurosci., vol. 27, no. 9, pp. 2349–2356, Feb. 2007, doi: 10.1523/JNEUROSCI.5587-06.2007.

[38] W. Gao et al., “Functional Network Development During the First Year: Relative Sequence and Socioeconomic Correlations,” Cereb. Cortex N. Y. NY, vol. 25, no. 9, pp. 2919–2928, Sep. 2015, doi: 10.1093/cercor/bhu088.

[39] L. Q. Uddin et al., “Salience network-based classification and prediction of symptom severity in children with autism,” JAMA Psychiatry, vol. 70, no. 8, pp. 869–879, Aug. 2013, doi: 10.1001/jamapsychiatry.2013.104.

[40] M. Attanasio, M. Mazza, I. Le Donne, A. Nigri, and M. Valenti, “Salience Network in Autism: preliminary results on functional connectivity analysis in resting state,” Eur. Arch. Psychiatry Clin. Neurosci., Dec. 2024, doi: 10.1007/s00406-024-01949-y.

[41] J. B. Girault et al., “Functional connectivity between the visual and salience networks and autistic social features at school-age,” J. Neurodev. Disord., vol. 17, no. 1, p. 23, Apr. 2025, doi: 10.1186/s11689-025-09613-9.

[42] S. A. Green, L. Hernandez, S. Y. Bookheimer, and M. Dapretto, “Salience Network Connectivity in Autism Is Related to Brain and Behavioral Markers of Sensory Overresponsivity,” J. Am. Acad. Child Adolesc. Psychiatry, vol. 55, no. 7, pp. 618–626.e1, Jul. 2016, doi: 10.1016/j.jaac.2016.04.013.

[43] K. K. Cummings et al., “Sex Differences in Salience Network Connectivity and its Relationship to Sensory Over-Responsivity in Youth with Autism Spectrum Disorder,” Autism Res., vol. 13, no. 9, pp. 1489–1500, 2020, doi: 10.1002/aur.2351.

[44] T. Tsang et al., “Salience network connectivity is altered in 6-week-old infants at heightened likelihood for developing autism,” Commun. Biol., vol. 7, no. 1, pp. 1–10, Apr. 2024, doi: 10.1038/s42003-024-06016-9.

[45] D. Scheinost et al., “Hypoconnectivity between anterior insula and amygdala associates with future vulnerabilities in social development in a neurodiverse sample of neonates,” Sci. Rep., vol. 12, p. 16230, Sep. 2022, doi: 10.1038/s41598-022-20617-6.

[46] J. Liu et al., “Atypical functional connectivity between the amygdala and visual, salience regions in infants with genetic liability for autism,” Cereb. Cortex, vol. 34, no. 13, pp. 30–39, May 2024, doi: 10.1093/cercor/bhae092.

[47] S. Ozonoff et al., “Familial Recurrence of Autism: Updates From the Baby Siblings Research Consortium,” Pediatrics, vol. 154, no. 2, p. e2023065297, Aug. 2024, doi: 10.1542/peds.2023-065297.

[48] J. J. Wolff et al., “Differences in white matter fiber tract development present from 6 to 24 months in infants with autism,” Am. J. Psychiatry, vol. 169, no. 6, pp. 589–600, Jun. 2012, doi: 10.1176/appi.ajp.2011.11091447.

[49] H. C. Hazlett et al., “Early brain development in infants at high risk for autism spectrum disorder,” Nature, vol. 542, no. 7641, pp. 348–351, Feb. 2017, doi: 10.1038/nature21369.

[50] J. Ciarrusta et al., “Emerging functional connectivity differences in newborn infants vulnerable to autism spectrum disorders,” Transl. Psychiatry, vol. 10, no. 1, pp. 1–10, May 2020, doi: 10.1038/s41398-020-0805-y.

[51] R. W. Emerson et al., “Functional neuroimaging of high-risk 6-month-old infants predicts a diagnosis of autism at 24 months of age,” Sci. Transl. Med., vol. 9, no. 393, p. eaag2882, Jun. 2017, doi: 10.1126/scitranslmed.aag2882.

[52] A. Nair et al., “Altered Thalamocortical Connectivity in 6-Week-Old Infants at High Familial Risk for Autism Spectrum Disorder,” Cereb. Cortex N. Y. N 1991, vol. 31, no. 9, pp. 4191–4205, Jul. 2021, doi: 10.1093/cercor/bhab078.

[53] J. R. Pruett Jr., et al., “Brain functional connectivity correlates of autism diagnosis and familial liability in 24-month-olds,” J. Neurodev. Disord., vol. 17, no. 1, p. 40, Jul. 2025, doi: 10.1186/s11689-025-09621-9.

[54] J. T. Elison et al., “White matter microstructure and atypical visual orienting in 7-month-olds at risk for autism,” Am. J. Psychiatry, vol. 170, no. 8, pp. 899–908, Aug. 2013, doi: 10.1176/appi.ajp.2012.12091150.

[55] J. J. Wolff et al., “Neural circuitry at age 6 months associated with later repetitive behavior and sensory responsiveness in autism,” Mol. Autism, vol. 8, no. 1, p. 8, Mar. 2017, doi: 10.1186/s13229-017-0126-z.

[56] A. T. Eggebrecht et al., “Joint Attention and Brain Functional Connectivity in Infants and Toddlers,” Cereb. Cortex N. Y. N 1991, vol. 27, no. 3, pp. 1709–1720, Mar. 2017, doi: 10.1093/cercor/bhw403.

[57] C. J. McKinnon et al., “Restricted and Repetitive Behavior and Brain Functional Connectivity in Infants at Risk for Developing Autism Spectrum Disorder,” Biol. Psychiatry Cogn. Neurosci. Neuroimaging, vol. 4, no. 1, pp. 50–61, Jan. 2019, doi: 10.1016/j.bpsc.2018.09.008.

[58] J. Liu et al., “Emerging atypicalities in functional connectivity of language-related networks in young infants at high familial risk for ASD,” Dev. Cogn. Neurosci., vol. 45, p. 100814, Oct. 2020, doi: 10.1016/j.dcn.2020.100814.

[59] L. Wagner et al., “Associations between thalamocortical functional connectivity and sensory over-responsivity in infants at high likelihood for ASD,” Cereb. Cortex, vol. 33, no. 12, pp. 8075–8086, Jun. 2023, doi: 10.1093/cercor/bhad100.

[60] E. R. Coco et al., “Sleep in Infants with Down Syndrome or Familial Likelihood of Autism in the First Year of Life,” J. Autism Dev. Disord., Jun. 2025, doi: 10.1007/s10803-025-06927-4.

[61] A. Klin, D. J. Lin, P. Gorrindo, G. Ramsay, and W. Jones, “Two-year-olds with autism orient to nonsocial contingencies rather than biological motion,” Nature, vol. 459, no. 7244, pp. 257–261, May 2009, doi: 10.1038/nature07868.

[62] A. J. Khan, A. Nair, C. L. Keown, M. C. Datko, A. J. Lincoln, and R.-A. Müller, “Cerebro-cerebellar Resting-State Functional Connectivity in Children and Adolescents with Autism Spectrum Disorder,” Biol. Psychiatry, vol. 78, no. 9, pp. 625–634, Nov. 2015, doi: 10.1016/j.biopsych.2015.03.024.

[63] K. M. Igelström, T. W. Webb, and M. S. A. Graziano, “Functional Connectivity Between the Temporoparietal Cortex and Cerebellum in Autism Spectrum Disorder,” Cereb. Cortex, vol. 27, no. 4, pp. 2617–2627, Apr. 2017, doi: 10.1093/cercor/bhw079.

[64] T. C. Ramos, J. B. Balardin, J. R. Sato, and A. Fujita, “Abnormal Cortico-Cerebellar Functional Connectivity in Autism Spectrum Disorder,” Front. Syst. Neurosci., vol. 12, Jan. 2019, doi: 10.3389/fnsys.2018.00074.

[65] R. Hanaie et al., “Aberrant Cerebellar–Cerebral Functional Connectivity in Children and Adolescents With Autism Spectrum Disorder,” Front. Hum. Neurosci., vol. 12, Nov. 2018, doi: 10.3389/fnhum.2018.00454.

[66] R. E. W. Smith, J. A. Avery, G. L. Wallace, L. Kenworthy, S. J. Gotts, and A. Martin, “Sex Differences in Resting-State Functional Connectivity of the Cerebellum in Autism Spectrum Disorder,” Front. Hum. Neurosci., vol. 13, Apr. 2019, doi: 10.3389/fnhum.2019.00104.

[67] M. E. Cakar et al., “Functional connectivity of the sensorimotor cerebellum in autism: associations with sensory over-responsivity,” Front. Psychiatry, vol. 15, Mar. 2024, doi: 10.3389/fpsyt.2024.1337921.

[68] L. Wagner et al., “Beyond motor learning: Insights from infant magnetic resonance imaging on the critical role of the cerebellum in behavioral development,” Dev. Cogn. Neurosci., vol. 72, p. 101514, Apr. 2025, doi: 10.1016/j.dcn.2025.101514.

[69] N. J. Okada et al., “Atypical cerebellar functional connectivity at 9 months of age predicts delayed socio-communicative profiles in infants at high and low risk for autism,” J. Child Psychol. Psychiatry, vol. 63, no. 9, pp. 1002–1016, 2022, doi: 10.1111/jcpp.13555.

[70] Z. W. Hawks et al., “A Prospective Evaluation of Infant Cerebellar-Cerebral Functional Connectivity in Relation to Behavioral Development in Autism Spectrum Disorder,” Biol. Psychiatry Glob. Open Sci., vol. 3, no. 1, pp. 149–161, Jan. 2023, doi: 10.1016/j.bpsgos.2021.12.004.

[71] S. Tikoo et al., “The Evolving Cerebellar and Cerebello-cortical Functional Connectivity Architecture during Infancy,” J. Neurosci., vol. 45, no. 11, Mar. 2025, doi: 10.1523/JNEUROSCI.1209-24.2025.

[72] C. S. Herzmann, A. Z. Snyder, J. K. Kenley, C. E. Rogers, J. S. Shimony, and C. D. Smyser, “Cerebellar Functional Connectivity in Term- and Very Preterm-Born Infants,” Cereb. Cortex N. Y. N 1991, vol. 29, no. 3, pp. 1174–1184, Mar. 2019, doi: 10.1093/cercor/bhy023.

[73] J. A. Kipping, T. A. Tuan, M. V. Fortier, and A. Qiu, “Asynchronous Development of Cerebellar, Cerebello-Cortical, and Cortico-Cortical Functional Networks in Infancy, Childhood, and Adulthood,” Cereb. Cortex N. Y. N 1991, vol. 27, no. 11, pp. 5170–5184, Nov. 2017, doi: 10.1093/cercor/bhw298.

[74] S. H. A. Chen and J. E. Desmond, “Cerebrocerebellar networks during articulatory rehearsal and verbal working memory tasks,” NeuroImage, vol. 24, no. 2, pp. 332–338, Jan. 2005, doi: 10.1016/j.neuroimage.2004.08.032.

[75] H. Nakatani, Y. Nakamura, and K. Okanoya, “Respective Involvement of the Right Cerebellar Crus I and II in Syntactic and Semantic Processing for Comprehension of Language,” Cerebellum Lond. Engl., vol. 22, no. 4, pp. 739–755, Aug. 2023, doi: 10.1007/s12311-022-01451-y.

[76] Y. Cheng et al., “Abnormal functional connectivity of the salience network in insomnia,” Brain Imaging Behav., vol. 16, no. 2, pp. 930–938, Apr. 2022, doi: 10.1007/s11682-021-00567-9.

[77] X. Yin, T. Jiang, Z. Song, L. Zhu, G. Wang, and J. Guo, “Increased functional connectivity within the salience network in patients with insomnia,” Sleep Breath., vol. 28, no. 3, pp. 1261–1271, Jun. 2024, doi: 10.1007/s11325-024-03002-7.

[78] C. B. Canto, Y. Onuki, B. Bruinsma, Y. D. van der Werf, and C. I. D. Zeeuw, “The Sleeping Cerebellum,” Trends Neurosci., vol. 40, no. 5, pp. 309–323, May 2017, doi: 10.1016/j.tins.2017.03.001.

[79] A. Jackson and W. Xu, “Role of cerebellum in sleep-dependent memory processes,” Front. Syst. Neurosci., vol. 17, Apr. 2023, doi: 10.3389/fnsys.2023.1154489.

[80] E. Benarroch, “What Is the Involvement of the Cerebellum During Sleep?,” Neurology, vol. 100, no. 12, pp. 572–577, Mar. 2023, doi: 10.1212/WNL.0000000000207161.

[81] C. Lord et al., “The Autism Diagnostic Observation Schedule—Generic: A Standard Measure of Social and Communication Deficits Associated with the Spectrum of Autism,” J. Autism Dev. Disord., vol. 30, no. 3, pp. 205–223, Jun. 2000, doi: 10.1023/A:1005592401947.

[82] C. Lord, M. Rutter, and A. Le Couteur, “Autism Diagnostic Interview-Revised: a revised version of a diagnostic interview for caregivers of individuals with possible pervasive developmental disorders,” J. Autism Dev. Disord., vol. 24, no. 5, pp. 659–685, Oct. 1994, doi: 10.1007/BF02172145.

[83] “Mullen EM. Mullen scales of early learning. Circle Pines, MN: AGS; 1995

[84] M. A. Gartstein and M. K. Rothbart, “Studying infant temperament via the Revised Infant Behavior Questionnaire,” Infant Behav. Dev., vol. 26, no. 1, pp. 64–86, Feb. 2003, doi: 10.1016/S0163-6383(02)00169-8.

[85] G. T. Baranek, F. J. David, M. D. Poe, W. L. Stone, and L. R. Watson, “Sensory Experiences Questionnaire: discriminating sensory features in young children with autism, developmental delays, and typical development,” J. Child Psychol. Psychiatry, vol. 47, no. 6, pp. 591–601, 2006, doi: 10.1111/j.1469-7610.2005.01546.x.

[86] V. D. Hohn, D. M. J. de Veld, K. J. S. Mataw, E. J. W. van Someren, and S. Begeer, “Insomnia Severity in Adults with Autism Spectrum Disorder is Associated with sensory Hyper-Reactivity and Social Skill Impairment,” J. Autism Dev. Disord., vol. 49, no. 5, pp. 2146–2155, May 2019, doi: 10.1007/s10803-019-03891-8.

[87] S. M. Smith et al., “Advances in functional and structural MR image analysis and implementation as FSL,” NeuroImage, vol. 23, pp. S208–S219, Jan. 2004, doi: 10.1016/j.neuroimage.2004.07.051.

[88] L. Chen et al., “A 4D infant brain volumetric atlas based on the UNC/UMN baby connectome project (BCP) cohort,” NeuroImage, vol. 253, p. 119097, Jun. 2022, doi: 10.1016/j.neuroimage.2022.119097.

[89] R. H. R. Pruim, M. Mennes, D. van Rooij, A. Llera, J. K. Buitelaar, and C. F. Beckmann, “ICA-AROMA: A robust ICA-based strategy for removing motion artifacts from fMRI data,” NeuroImage, vol. 112, pp. 267–277, May 2015, doi: 10.1016/j.neuroimage.2015.02.064.

[90] R. H. R. Pruim, M. Mennes, J. K. Buitelaar, and C. F. Beckmann, “Evaluation of ICA-AROMA and alternative strategies for motion artifact removal in resting state fMRI,” NeuroImage, vol. 112, pp. 278–287, May 2015, doi: 10.1016/j.neuroimage.2015.02.063.

[91] D. Carone, R. Licenik, S. Suri, L. Griffanti, N. Filippini, and J. Kennedy, “Impact of automated ICA-based denoising of fMRI data in acute stroke patients,” NeuroImage Clin., vol. 16, pp. 23–31, 2017, doi: 10.1016/j.nicl.2017.06.033.

[92] J. D. Power, A. Mitra, T. O. Laumann, A. Z. Snyder, B. L. Schlaggar, and S. E. Petersen, “Methods to detect, characterize, and remove motion artifact in resting state fMRI,” NeuroImage, vol. 84, pp. 320–341, Jan. 2014, doi: 10.1016/j.neuroimage.2013.08.048.

[93] Q. Luo et al., “Effective connectivity of the right anterior insula in schizophrenia: The salience network and task-negative to task-positive transition,” NeuroImage Clin., vol. 28, p. 102377, Jan. 2020, doi: 10.1016/j.nicl.2020.102377.

[94] J. X. O’Reilly, C. F. Beckmann, V. Tomassini, N. Ramnani, and H. Johansen-Berg, “Distinct and Overlapping Functional Zones in the Cerebellum Defined by Resting State Functional Connectivity,” Cereb. Cortex, vol. 20, no. 4, pp. 953–965, Apr. 2010, doi: 10.1093/cercor/bhp157.

[95] A. M. D’Mello and C. J. Stoodley, “Cerebro-cerebellar circuits in autism spectrum disorder,” Front. Neurosci., vol. 9, Nov. 2015, doi: 10.3389/fnins.2015.00408.

[96] M. King, C. R. Hernandez-Castillo, R. A. Poldrack, R. B. Ivry, and J. Diedrichsen, “Functional boundaries in the human cerebellum revealed by a multi-domain task battery,” Nat. Neurosci., vol. 22, no. 8, pp. 1371–1378, Aug. 2019, doi: 10.1038/s41593-019-0436-x.

[97] M. Leggio and G. Olivito, “Chapter 5 - Topography of the cerebellum in relation to social brain regions and emotions,” in Handbook of Clinical Neurology, vol. 154, M. Manto and T. A. G. M. Huisman, Eds., in The Cerebellum: From Embryology to Diagnostic Investigations, vol. 154., Elsevier, 2018, pp. 71–84. doi: 10.1016/B978-0-444-63956-1.00005-9.

[98] F. Shi et al., “Infant Brain Atlases from Neonates to 1- and 2-Year-Olds,” PLOS ONE, vol. 6, no. 4, p. e18746, Apr. 2011, doi: 10.1371/journal.pone.0018746.

[99] J. Diedrichsen, J. H. Balsters, J. Flavell, E. Cussans, and N. Ramnani, “A probabilistic MR atlas of the human cerebellum,” NeuroImage, vol. 46, no. 1, pp. 39–46, May 2009, doi: 10.1016/j.neuroimage.2009.01.045.

[100] E. Mahmoudi, N. Mirzakhany, S. M. Tabatabaee, S. Fallah, and M. Shahbazi, “The Relationship between Sensory Processing Disorder and Quality of Sleep in Children with Autism Spectrum Disorder and Learning Disorder from 6 to 14 Years’ Old,” J. Clin. Physiother. Res., vol. 4, no. 3, Art. no. 3, Jul. 2019, doi: 10.22037/jcpr.v4i3.29465.

[101] G. T. Baranek et al., “Cascading effects of attention disengagement and sensory seeking on social symptoms in a community sample of infants at-risk for a future diagnosis of autism spectrum disorder,” Dev. Cogn. Neurosci., vol. 29, pp. 30–40, Jan. 2018, doi: 10.1016/j.dcn.2017.08.006.

[102] R. Grzadzinski et al., “Pre-symptomatic intervention for autism spectrum disorder (ASD): defining a research agenda,” J. Neurodev. Disord., vol. 13, no. 1, p. 49, Oct. 2021, doi: 10.1186/s11689-021-09393-y.

[103] Y.-J. Chen, J. Sideris, L. R. Watson, E. R. Crais, and G. T. Baranek, “Early developmental profiles of sensory features and links to school-age adaptive and maladaptive outcomes: A birth cohort investigation,” Dev. Psychopathol., vol. 36, no. 1, pp. 291–301, Feb. 2024, doi: 10.1017/S0954579422001195.

[104] T. Charman et al., “Non-ASD outcomes at 36 months in siblings at familial risk for autism spectrum disorder (ASD): A baby siblings research consortium (BSRC) study,” Autism Res. Off. J. Int. Soc. Autism Res., vol. 10, no. 1, pp. 169–178, Jan. 2017, doi: 10.1002/aur.1669.

[105] L. Yi, Q. Wang, C. Song, and Z. R. Han, “Hypo- or hyperarousal? The mechanisms underlying social information processing in autism,” Child Dev. Perspect., vol. 16, no. 4, pp. 215–222, 2022, doi: 10.1111/cdep.12466.

[106] D. Riemann et al., “The hyperarousal model of insomnia: A review of the concept and its evidence,” Sleep Med. Rev., vol. 14, no. 1, pp. 19–31, Feb. 2010, doi: 10.1016/j.smrv.2009.04.002.

[107] C. Habas et al., “Distinct Cerebellar Contributions to Intrinsic Connectivity Networks,” J. Neurosci., vol. 29, no. 26, pp. 8586–8594, Jul. 2009, doi: 10.1523/JNEUROSCI.1868-09.2009.

[108] L. S. Popa and T. J. Ebner, “Cerebellum, Predictions and Errors,” Front. Cell. Neurosci., vol. 12, Jan. 2019, doi: 10.3389/fncel.2018.00524.

[109] J.-W. Jeong, V. N. Tiwari, M. E. Behen, H. T. Chugani, and D. C. Chugani, “In vivo detection of reduced Purkinje cell fibers with diffusion MRI tractography in children with autistic spectrum disorders,” Front. Hum. Neurosci., vol. 8, Feb. 2014, doi: 10.3389/fnhum.2014.00110.

[110] M. Verly et al., “Altered functional connectivity of the language network in ASD: Role of classical language areas and cerebellum,” NeuroImage Clin., vol. 4, pp. 374–382, Jan. 2014, doi: 10.1016/j.nicl.2014.01.008.

[111] A. M. D’Mello, D. Crocetti, S. H. Mostofsky, and C. J. Stoodley, “Cerebellar gray matter and lobular volumes correlate with core autism symptoms,” NeuroImage Clin., vol. 7, pp. 631–639, Jan. 2015, doi: 10.1016/j.nicl.2015.02.007.

[112] F. d’Oleire Uquillas et al., “Multimodal evidence for cerebellar influence on cortical development in autism: structural growth amidst functional disruption,” Mol. Psychiatry, vol. 30, no. 4, pp. 1558–1572, Apr. 2025, doi: 10.1038/s41380-024-02769-1.

[113] E. Sefik et al., “Structural deviations of the posterior fossa and the cerebellum and their cognitive links in a neurodevelopmental deletion syndrome,” Mol. Psychiatry, vol. 29, no. 11, pp. 3395–3411, 2024, doi: 10.1038/s41380-024-02584-8.

[114] J. A. Kipping, W. Grodd, V. Kumar, M. Taubert, A. Villringer, and D. S. Margulies, “Overlapping and parallel cerebello-cerebral networks contributing to sensorimotor control: An intrinsic functional connectivity study,” NeuroImage, vol. 83, pp. 837–848, Dec. 2013, doi: 10.1016/j.neuroimage.2013.07.027.

[115] G. P. D. Argyropoulos, “The cerebellum, internal models and prediction in ‘non-motor’ aspects of language: A critical review,” Brain Lang., vol. 161, pp. 4–17, Oct. 2016, doi: 10.1016/j.bandl.2015.08.003.

[116] A. A. Sokolov, R. C. Miall, and R. B. Ivry, “The Cerebellum: Adaptive Prediction for Movement and Cognition,” Trends Cogn. Sci., vol. 21, no. 5, pp. 313–332, May 2017, doi: 10.1016/j.tics.2017.02.005.

[117] A. M. Berlijn et al., “The Role of the Human Cerebellum for Learning from and Processing of External Feedback in Non-Motor Learning: A Systematic Review,” The Cerebellum, vol. 23, no. 4, pp. 1532–1551, Aug. 2024, doi: 10.1007/s12311-024-01669-y.

[118] J. Ciarrusta et al., “Social Brain Functional Maturation in Newborn Infants With and Without a Family History of Autism Spectrum Disorder,” JAMA Netw. Open, vol. 2, no. 4, p. e191868, Apr. 2019, doi: 10.1001/jamanetworkopen.2019.1868.

[119] O. J. Veatch, A. C. Maxwell-Horn, and B. A. Malow, “Sleep in Autism Spectrum Disorders,” Curr. Sleep Med. Rep., vol. 1, no. 2, pp. 131–140, Jun. 2015, doi: 10.1007/s40675-015-0012-1.

[120] G. Alrousan, A. Hassan, A. A. Pillai, F. Atrooz, and S. Salim, “Early Life Sleep Deprivation and Brain Development: Insights From Human and Animal Studies,” Front. Neurosci., vol. 16, May 2022, doi: 10.3389/fnins.2022.833786.

[121] L. Zwaigenbaum et al., “Early Intervention for Children With Autism Spectrum Disorder Under 3 Years of Age: Recommendations for Practice and Research,” Pediatrics, vol. 136 Suppl 1, no. Suppl 1, pp. S60–81, Oct. 2015, doi: 10.1542/peds.2014-3667E.

[122] E. A. Fuller and A. P. Kaiser, “The Effects of Early Intervention on Social Communication Outcomes for Children with Autism Spectrum Disorder: A Meta-analysis,” J. Autism Dev. Disord., vol. 50, no. 5, pp. 1683–1700, May 2020, doi: 10.1007/s10803-019-03927-z.

[123] L. E. Miller, K. A. Perkins, Y. G. Dai, and D. A. Fein, “Comparison of parent report and direct assessment of child skills in toddlers,” Res. Autism Spectr. Disord., vol. 41-42, pp. 57–65, Sep. 2017, doi: 10.1016/j.rasd.2017.08.002.

[124] S. F. Schoch, S. Kurth, and H. Werner, “Actigraphy in sleep research with infants and young children: Current practices and future benefits of standardized reporting,” J. Sleep Res., vol. 30, no. 3, p. e13134, Jun. 2021, doi: 10.1111/jsr.13134.

[125] P. M. Siper, A. Kolevzon, A. T. Wang, J. D. Buxbaum, and T. Tavassoli, “A clinician-administered observation and corresponding caregiver interview capturing DSM-5 sensory reactivity symptoms in children with ASD,” Autism Res. Off. J. Int. Soc. Autism Res., vol. 10, no. 6, pp. 1133–1140, Jun. 2017, doi: 10.1002/aur.1750.

[126] A. Ben-Sasson, E. Gal, R. Fluss, N. Katz-Zetler, and S. A. Cermak, “Update of a Meta-analysis of Sensory Symptoms in ASD: A New Decade of Research,” J. Autism Dev. Disord., vol. 49, no. 12, pp. 4974–4996, Dec. 2019, doi: 10.1007/s10803-019-04180-0.

[127] M. Korom et al., “Dear reviewers: Responses to common reviewer critiques about infant neuroimaging studies,” Dev. Cogn. Neurosci., vol. 53, p. 101055, Feb. 2022, doi: 10.1016/j.dcn.2021.101055.

