## Supplemental Materials for "Salience Network Connectivity Relates to Sleep and Sensory Over-Responsivity in Infants at High and Low Likelihood for Autism"

**Supplementary Material**

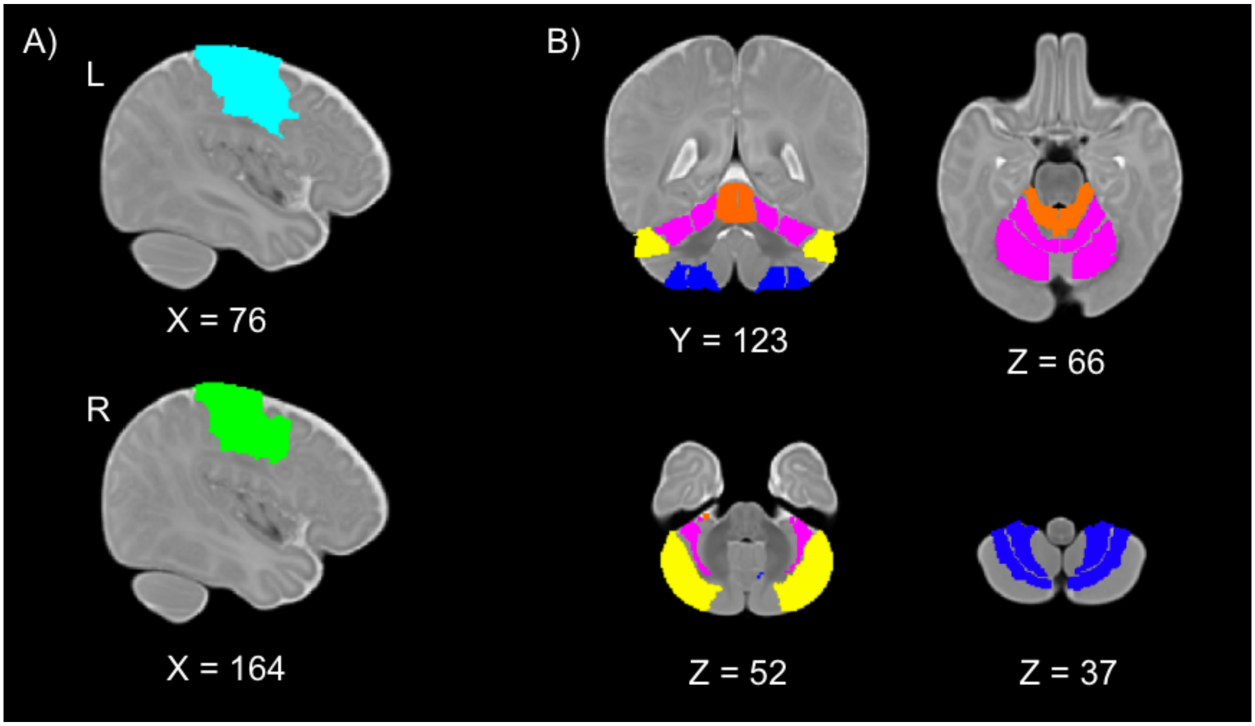


**Supplementary Figure S1. ROI masks used for between-group comparisons and behavioral regression analyses.** Left sensorimotor cortex (light blue), right sensorimotor cortex (green), cerebellum crus I (yellow), cerebellum lobule I-IV (orange), cerebellum lobule V-VI (magenta), and cerebellum lobule VIIIA/B (dark blue).

| **When going to sleep at night, how often did your baby:** | |
|---|---|
| 21. Fall asleep within 10 minutes? | R |
| 22. Have a hard time settling down to sleep? | |
| 23. Settle down to sleep easily? | R |
| **When your baby awoke at night, how often did s/he:** | |
| 24. Have a hard time going back to sleep? | |
| 25. Go back to sleep immediately? | R |

**Supplementary Table S1. Infant Sleep-Onset Problems (ISOP) score**. Five questions from the Infant Behavior Questionnaire (collected at 6 and 12 months) were used to generate the Infant Sleep-Onset Problems (ISOP) score. Questions were scored on a 7-point scale ranging from “never” to “always”. R indicates reverse-scored items. Higher scores indicate worse sleep-onset problems.

| |
|---|
| **Experiences with Sound:** |
| 1. Does your child react sensitively or startle easily to unexpected or loud sounds?<br>5. Does your child notice sounds in the environment (such as planes, trains, faucets dripping, lights buzzing, etc.) before other people do?<br>6. Does your child show distress (startles, covers ears, etc.) during loud conversations or singing? |
| **Experiences with Sight:** |
| 8. Is your child disturbed by too much light inside or brightness outside?<br>11. Does your child avoid looking at your face during social games/play? |
| **Experiences with Touch:** |
| 14. Does your child dislike cuddling or being held?<br>15. Does your child show distress during grooming? (For example: cries or fusses during face washing, hair combing, fingernail cutting, or teeth brushing.)<br>16. Does your child avoid touching certain textures (such as fuzzy or squishy toys) or playing with messy materials (such as sand, lotion)?<br>17. Does your child react negatively or pull away when touched by a person? (For example: pulls away when head is patted.)<br>18. Does your child have trouble adjusting to the water temperature during bath time for does he/she dislike being in water?<br>20. Does your child dislike being tickled? |

**Supplementary Table S2. Sensory Over-Responsivity (SOR) score**. Eleven questions from the Sensory Experiences Questionnaire (collected at 12 and 24 months) were used to generate the Sensory Over-Responsivity (SOR) score. Questions were scored on a 5-point scale ranging from “Almost Never” to “Almost Always.” Higher scores indicate greater frequency or intensity of sensory features.
